# Neurobehavioral Profiles of Inhibitory-Control Stratify Vulnerability and Resilience under Childhood Poverty

**DOI:** 10.64898/2026.04.18.26350994

**Authors:** Bowen Hu, Tongxi Yang, Yuanyuan Hu, Moyu Liu, Shuping Tan, Xinying Li, Shaozheng Qin

## Abstract

**Objective:** Childhood poverty is a high-risk context that involves diverse adversities, making it difficult to understand how poverty confers later psychopathology risk and why some children remain resilient despite growing up in poverty. To address this heterogeneity, we quantified adversity-linked vulnerability as adversity-psychopathology coupling and tested whether childhood poverty amplifies this coupling and whether multilevel inhibitory-control profiles stratify vulnerability and resilience within poverty-exposed youth.

**Methods:** We analyzed 10,112 youth (48.4% female; mean age = 9.92 years) from the Adolescent Brain Cognitive Development Study, linking baseline cumulative early-life adversity (ELA) to later behavioral problems across 4 waves. In the stop-signal task fMRI subsample of 7,401 youth, semi-supervised clustering of inhibitory-control activation identified neurofunctional subtypes within poverty-exposed youth. We also tested temperamental inhibitory control as an additional moderator.

**Results:** Childhood poverty amplified the association between cumulative ELA and behavioral problems at baseline (Δβ = 0.088; *P* < .001) and across follow-up waves. Two neurofunctional subtypes were identified within poverty-exposed youth: subtype-1 showed greater vulnerability than higher-income peers (Δβ = 0.149; *P* < .001), whereas subtype-2 showed attenuated vulnerability and did not differ from higher-income peers (Δβ = 0.049; *P* = .135); this pattern persisted longitudinally. Among poverty-exposed youth in subtype-2 with high temperamental inhibitory control, the association between cumulative ELA and later behavioral problems was no longer significant.

**Conclusions:** Childhood poverty strengthened the translation of adversity burden into later behavioral problems, but inhibitory-control profiles differentiated higher- and lower-risk pathways within poverty, highlighting inhibitory control as a candidate target for prevention.

## Introduction

Childhood poverty is a complex high-risk context for mental health.^1^ It involves greater exposure to early-life adversities (ELAs) and chronic strain that may shape neurocognitive development.^2,3^ Yet outcomes among poverty-exposed youth are highly heterogeneous. Many show elevated behavioral problems, whereas others show attenuated vulnerability or relative resilience.^4^ Explaining why similarly exposed youth diverge into heightened vulnerability versus relative resilience remains a central challenge for developmental psychopathology and prevention design.

A useful way to characterize this process is to quantify adversity-linked vulnerability, or adversity–psychopathology coupling, as the dose–response slope linking incremental adversity burden to incremental behavioral problems.^5,6^ If poverty amplifies vulnerability, then at the same level of cumulative ELA, poverty-exposed youth should show steeper increases in behavioral problems than higher-income peers. This slope-based approach distinguishes baseline symptom burden from symptom sensitivity to accumulating adversity, thereby capturing elevated risk even when the differences in average symptom levels are modest.^6,7^

However, vulnerability-based risk is unlikely to be homogeneous within poverty. Poverty bundles heterogeneous stressors, resources, and coping opportunities, and children differ in self-regulatory capacity and developmental context.^2,8^ Accordingly, poverty may diversify the pathways linking adversity to psychopathology.^4,9^ Empirically, this implies that poverty-exposed youth may comprise distinct subgroups with divergent adversity–psychopathology coupling, motivating multivariate approaches that can stratify vulnerability and resilience within poverty.^10,11^

Inhibitory control is a strong candidate process for distinguishing vulnerability and resilience within poverty. It supports goal-directed behavior and emotion regulation and undergoes protracted development. It is also sensitive to stress-related inputs that calibrate regulatory systems across development.^12,13^ At the neurocognitive level, inhibitory control depends on distributed control circuitry that shows marked individual differences in task-evoked recruitment.^14^ In parallel, temperamental inhibitory control captures individual differences in everyday self-regulation.^15^ These indices may diverge under stress because task activation can reflect compensatory recruitment under higher demands, whereas temperament indexes stable self-regulatory style across contexts.^13,16^ A multilevel phenotype that integrates activation patterns with temperament is therefore needed to distinguish vulnerability from resilience within poverty.

Here we use the Adolescent Brain Cognitive Development Study to test a neurobehavioral account of vulnerability and resilience under childhood poverty.^17^ We first examine whether childhood poverty amplifies the translation of cumulative ELA into later behavioral problems across late childhood and adolescence. We then apply heterogeneity-aware subtyping to multivariate stop-signal task–evoked activation to identify neurofunctional subtypes associated with divergent vulnerability and resilience profiles within poverty-exposed youth.^11^ Next, we test whether temperamental inhibitory control further sharpens these subtype-defined profiles. Finally, to biologically contextualize subtype patterns, we additionally perform exploratory multiscale mapping to neurotransmitter receptor distributions and cortical gene-expression gradients.^18^

## Methods

### Sample

We conducted a prospective cohort analysis of the Adolescent Brain Cognitive Development (ABCD, release 5.1) Study, a community-based US cohort recruited from 21 sites with repeated assessments from late childhood into adolescence. The ABCD protocol was approved by the institutional review boards at each site. Parents or legal guardians provided written informed consent and children provided assent. Sex at birth, age, race and ethnicity were reported by parents or guardians at wave-1 using ABCD standard categories to characterize the sample demographics. Because gender identity was not analyzed in this study, we report caregiver-reported sex assigned at birth. This study is reported in accordance with the Strengthening the Reporting of Observational Studies in Epidemiology (STROBE) reporting guideline.

The analytic sample included participants with complete baseline demographic information and at least 1 assessment of behavioral problems (CBCL Total Problems) across study waves. Poverty exposure was defined at wave-1 using the federal poverty threshold based on household income and family size; children above the threshold served as the higher-income comparison group. Primary analyses included 10,112 participants at wave-1, with CBCL data available for 9,624 at wave-2, 9,394 at wave-3, and 8,759 at wave-4; HYDRA subtyping analyses included 7,401 participants with usable stop-signal task fMRI data at wave-1, with CBCL data available for 7,087 at wave-2, 6,927 at wave-3, and 6,500 at wave-4. Additional cohort selection and imaging quality-control procedures are provided in Supplementary Methods 1 and 4. Sample size was determined by the number of ABCD participants who met prespecified eligibility criteria for this secondary analysis; no a priori power calculation was performed.

### Early-Life Adversity (ELA) Burden

We operationalized ELA using 14 standardized indicators spanning prenatal exposure, child health/trauma, family functioning, peer adversity, and school- and neighborhood-level contexts. Missing values on individual ELA indicators were mean-imputed (Supplementary Methods 2). Cumulative ELA was computed by rescaling each indicator to 0–1 and summing them. The primary index used 8 indicators showing nominal poverty amplification, and sensitivity analyses using all 14 indicators and complete-case estimation yielded convergent results (Supplementary Results 3). We treated this 8-ELA index as a parsimonious, data-enriched summary of poverty-sensitive adversity burden; importantly, convergent findings with the full 14-ELA index indicated that the main conclusions did not depend on this selection.

### Measurements and Neurobehavioral Moderators

Behavioral problems were indexed by the Child Behavior Checklist (CBCL) Total Problems scale (age- and sex-normed T scores) at baseline and follow-up waves.^19^ Temperamental inhibitory control was assessed at wave-3 using the caregiver-reported inhibitory control subscale of the Early Adolescent Temperament Questionnaire–Revised (EATQ-R).^20^ Inhibitory-control neurofunction was indexed by stop-signal task (SST)–evoked activation.^21^ SST–evoked activation was quantified as region-of-interest (ROI)–level β estimates for the correct stop vs correct go contrast. ROIs comprised 148 Destrieux cortical parcels and 30 ASEG subcortical regions (178 total), as derived from the ABCD pipeline.^21,22^ Additional details regarding measurements, neuroimaging preprocessing, ROI extraction, and quality control are provided in Supplementary Methods 3 and 4.

### Neurofunctional Subtyping of Inhibitory-Control Activation

To identify within-poverty heterogeneity in inhibitory-control neurofunction, we applied HYDRA (Heterogeneity Through Discriminative Analysis), a semi-supervised discriminative clustering approach that derives subtypes among poverty-exposed youth relative to higher-income youth.^11^ Features were 178 ROI-level activation estimates (correct stop > correct go) spanning cortical and subcortical regions. Prior to model fitting, activation features were residualized for age, sex at birth, race and ethnicity, site, and mean framewise displacement. The final 2-subtype solution was selected based on prespecified stability testing (Supplementary Methods 5).

### Statistical Analyses

Adversity-linked vulnerability was quantified as the association (slope) between cumulative ELA burden and CBCL Total Problems using linear mixed-effects models with random intercepts for site and family, and (for longitudinal analyses) an additional random intercept for participant (Supplementary Methods 6). Primary tests evaluated effect modification by poverty status in the full cohort and, within the SST fMRI subset, by a 3-level group factor (higher-income, poverty subtype 1, poverty subtype 2), including moderation by temperamental inhibitory control (Supplementary Methods 6). All models adjusted for age, sex, and race/ethnicity, and log-transformed income-to-needs ratio; longitudinal models additionally included wave and age at visit. We controlled the false discovery rate within families of related tests (Benjamini-Hochberg; reported as *P*_FDR_). Prespecified sensitivity analyses examined alternative cumulative-ELA construction, complete-case estimation without imputation, and one-child-per-family reanalyses (Supplementary Methods 7).

### Exploratory Molecular Mapping

We performed exploratory molecular mapping to provide biological context for subtype activation patterns using PET receptor/transporter atlases and AHBA transcriptomic data (Supplementary Methods 8–10).^18^

## Results

### Sample Characteristics

The sample included 10,112 children with complete demographic data (mean [SD] age = 9.92 [0.63] years; 48.40% female; 55.40% White). Poverty-exposed youth comprised 1,492 participants (14.80%). At baseline, poverty-exposed youth showed higher cumulative ELA (*P* < .001; Cohen’s *d* = 0.85) and higher behavioral problems (CBCL Total Problems; *P* < .001; Cohen’s *d* = 0.26) than higher-income youth (Table 1).

**Table 1.**
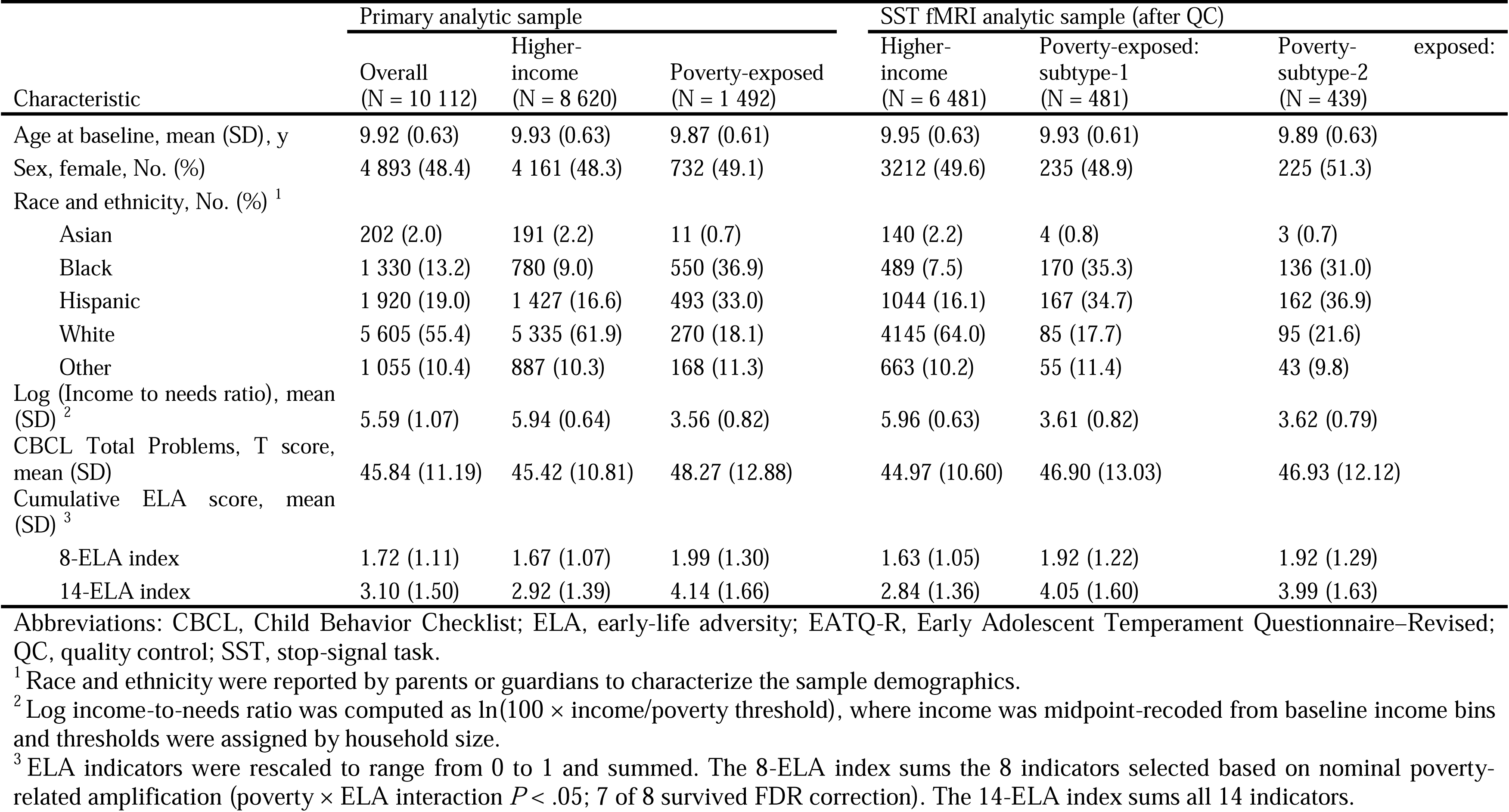
Baseline Characteristics of Study Participants.

### Poverty and Adversity-Linked Vulnerability

Across late childhood into adolescence, poverty exposure amplified the association between cumulative ELA burden and behavioral problems, indicating steeper adversity–psychopathology coupling under poverty. Poverty significantly moderated 7 of 14 ELA-specific associations with CBCL Total Problems after FDR correction (*P*_FDR_ < .05; Figure 1A; Supplementary Table 1). Building on these ELA-specific findings, we defined cumulative ELA burden using the subset of ELAs with evidence of poverty-related amplification in ELA-specific models (8 of the 14 ELAs; *P* < .05, uncorrected for multiple comparisons), to form a parsimonious cumulative burden measure. At baseline, cumulative ELA was more strongly associated with CBCL Total Problems in poverty-exposed youth than in higher-income youth (Δβ = 0.088; SE = 0.020; *P* < .001; Figure 1B), and this amplification remained evident at follow-up waves (Figure 1C; e.g., at wave-4: Δβ = 0.061, SE = 0.022, *P*_FDR_ < .01; Supplementary Table 2 and 3).

**Figure 1.**
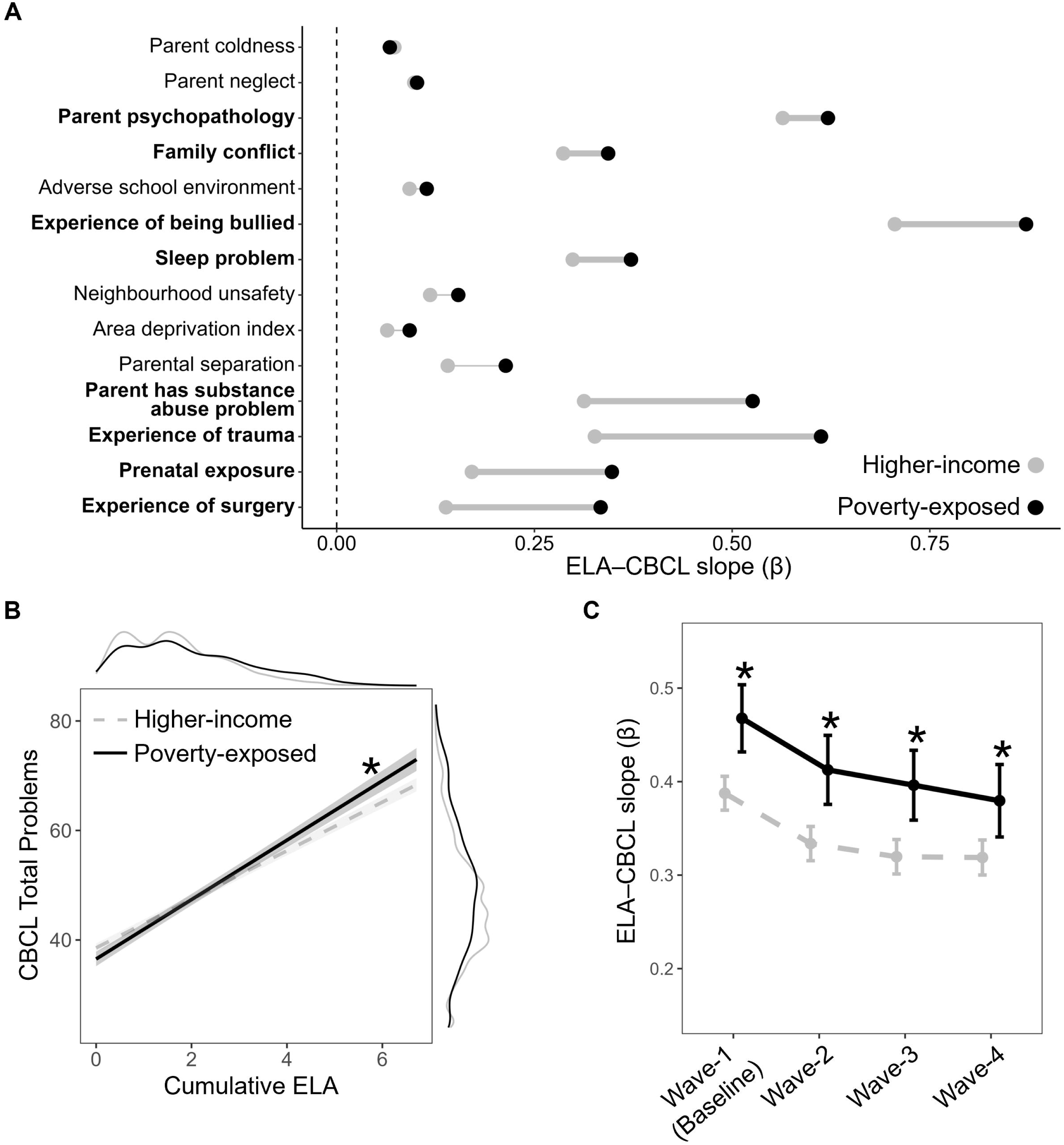
Poverty Exposure Amplifies Adversity–Psychopathology Coupling Across Late Childhood and Adolescence. **A,** ELA–CBCL slopes (β) from ELA-specific models relating each baseline ELA indicator to CBCL Total Problems, shown separately for higher-income and poverty-exposed youth. Points denote model-implied group-specific slopes; connecting lines facilitate visual comparison across groups, with thicker lines and bolded text indicating nominal evidence of poverty-related amplification (poverty × ELA interaction, *P* < .05). **B,** Baseline association between cumulative ELA burden (primary index) and CBCL Total Problems for higher-income and poverty-exposed youth. Lines depict model-implied associations; shaded bands represent 95% CIs. Marginal density plots show the distributions of cumulative ELA (top) and CBCL Total Problems (right) by group. Asterisk indicates a significant between-group difference in the ELA–CBCL slope (ELA × poverty interaction, *P* < .05). **C,** Wave-specific ELA–CBCL slopes (β) for the association between cumulative ELA burden and CBCL Total Problems derived from the longitudinal mixed-effects model (waves 1–4). Error bars represent 95% CIs. Asterisks indicate significant between-group differences in wave-specific ELA–CBCL slopes (group differences in estimated marginal ELA slopes at each wave, *P*_FDR_ < .05). Abbreviations: CBCL, Child Behavior Checklist; ELA, early-life adversity; CI, confidence interval. Alt text: Dot and line plots show ELA–CBCL slopes are higher in poverty-exposed youth, and CBCL rises more steeply with cumulative ELA.

### Neurofunctional Subtypes

HYDRA applied to multivariate SST–evoked activation identified two neurofunctional subtypes within poverty-exposed youth (Figure 2A). This 2-subtype solution was the most stable in cross-validation (ARI = 0.505) and outperformed label-shuffled null solutions (*P* < .05; Supplementary Methods 5). The subtypes diverged in distributed activation profiles relative to higher-income youth (Figure 2B and 2C; Supplementary Table 4). Subtype-1 exhibited higher activation across distributed regions, whereas subtype-2 exhibited lower activation. The two subtypes did not differ significantly in demographics, baseline ELA exposure, CBCL Total Problems scores, and cognitive performance (Supplementary Table 5); structural MRI differences did not survive multiple-comparison correction (Supplementary Results 1).

**Figure 2.**
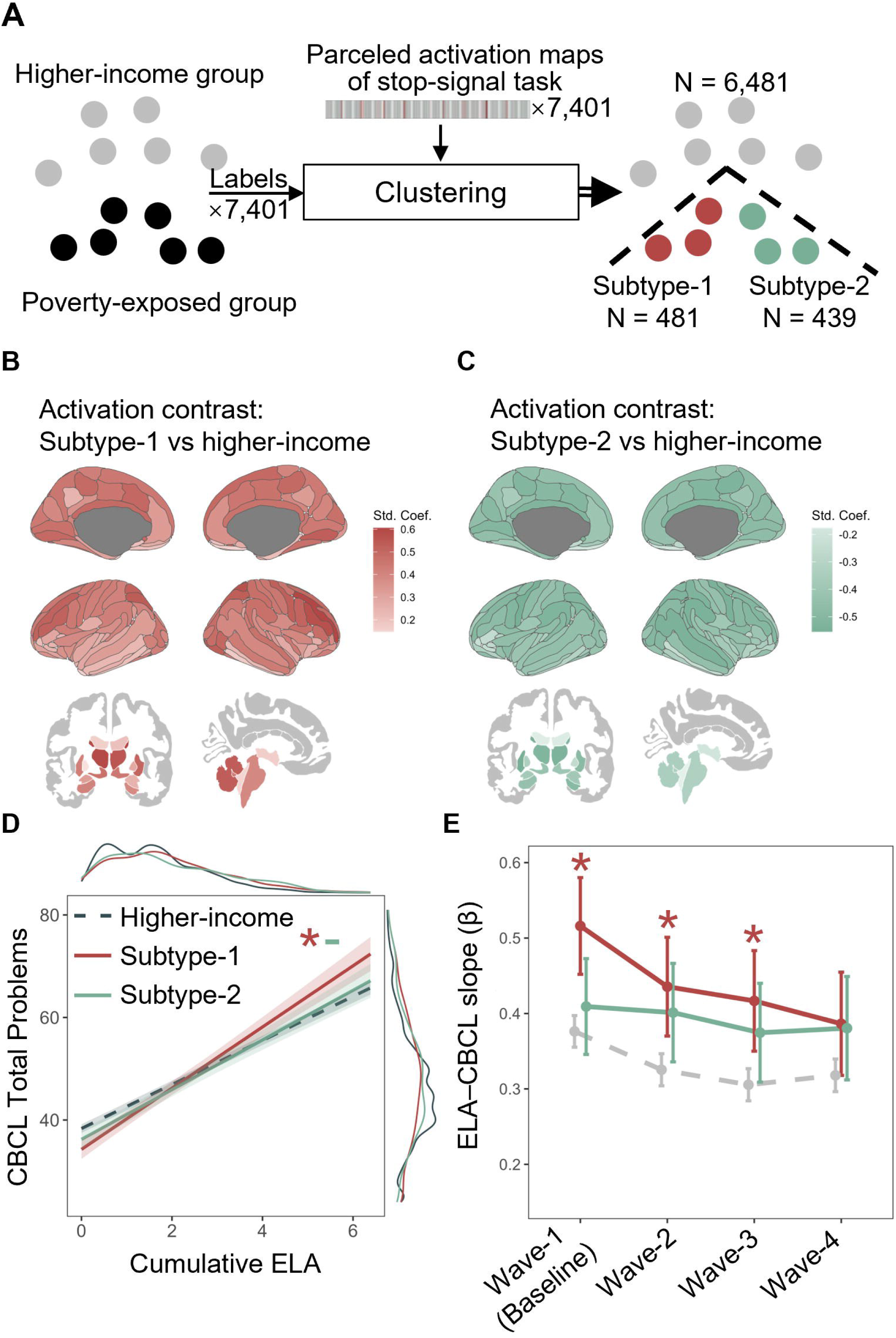
Neurofunctional Subtypes of Inhibitory-Control Activation Stratify Adversity–Psychopathology Coupling Within Poverty-Exposed Youth. **A,** Overview of HYDRA subtyping in the fMRI analytic sample. Parcellated stop-signal task activation estimates were used as features, and poverty status labels were used to derive neurofunctional subtypes among poverty-exposed youth relative to a higher-income comparison group. **B,** Parcel-wise activation contrasts for subtype-1 vs higher-income youth, displayed as standardized coefficients summarizing subtype differences in stop-signal task–evoked activation (correct stop vs correct go). Warmer colors indicate higher activation in subtype-1 relative to higher-income youth. **C,** Parcel-wise activation contrasts for subtype-2 vs higher-income youth, displayed as standardized coefficients. Cooler colors indicate lower activation in subtype-2 relative to higher-income youth. **D,** Baseline association between cumulative early-life adversity (ELA) burden and CBCL Total Problems for higher-income youth and the 2 poverty-exposed subtypes. Lines indicate model-implied associations; shaded bands represent 95% CIs. Marginal density plots show the distributions of cumulative ELA (top) and CBCL Total Problems (right) by group. **E,** Wave-specific ELA–CBCL slopes (β) for the association between cumulative ELA burden and CBCL Total Problems derived from the longitudinal mixed-effects model (waves 1–4). Error bars represent 95% CIs. Asterisks indicate significant between-group differences in wave-specific ELA–CBCL slopes (group differences in estimated marginal ELA slopes at each wave, subtypes vs high-income group, *P*_FDR_ < .05). Abbreviations: CBCL, Child Behavior Checklist; ELA, early-life adversity; fMRI, functional magnetic resonance imaging; CI, confidence interval. Alt text: Flowchart and brain maps show HYDRA-derived fMRI subtypes; subtype-1 hyperactivation and subtype-2 hypoactivation stratify ELA–CBCL coupling.

These subtypes defined distinct profiles within poverty-exposed youth. Subtype-1 showed greater vulnerability than the higher-income comparison group (Figure 2D), with cumulative ELA predicting CBCL Total Problems with β = 0.533 (SE = 0.032) in subtype-1 versus *β* = 0.384 (SE = 0.010) in higher-income youth (Δβ = 0.149; SE = 0.033; *P* < .001; Figure 2D). In contrast, subtype-2 (β = 0.433; SE = 0.031) showed attenuated vulnerability, with coupling comparable to the higher-income group (Δβ = 0.049; SE = 0.033; P = .135). Longitudinal contrasts showed that subtype-1 continued to show greater vulnerability than higher-income youth, whereas subtype-2 continued to show attenuated vulnerability (Figure 2E; Supplementary Table 2 and 6).

We also tested whether this attenuated-vulnerability pattern could be attributed to any single ROI. In ROI-wise models, no region showed a significant poverty × ELA × activation interaction after FDR correction, and nominal effects did not replicate prospectively (Supplementary Table 7), supporting a distributed, multivariate signature of vulnerability stratification. Stop-signal task performance (stop-signal reaction time) did not moderate ELA–CBCL coupling, nor did this moderation vary by poverty status (Supplementary Results 2).

### Temperamental Moderation

In the full sample, higher inhibitory-control temperament predicted weaker adversity-linked vulnerability at follow-up (cumulative ELA × temperament: wave-3, β = −0.030, SE = 0.008, *P* < .001; wave-4, β = −0.024, SE = 0.009, *P* = .005). This buffering of adversity-linked vulnerability tended to be stronger under poverty (poverty × cumulative ELA × temperament: wave-3, β = −0.039, SE = 0.020, *P* = .054; wave-4, β = −0.051, SE = 0.023, *P* = .024; simple slope analyses are in the Supplementary Table 8.1).

Temperament and neurofunctional subtype jointly sharpened subtype-defined vulnerability and resilience profiles within poverty (subtype × cumulative ELA × temperament: wave-3, F(2, 6590) = 3.360, *P* = .035; wave-4, F(2, 5829.4) = 3.017, *P* = .049; Figure 3A). This buffering emerged most strongly and selectively in subtype-2: among those with high temperament, cumulative ELA no longer predicted later behavioral problems at either follow-up (wave-3, β = 0.088, SE = 0.090, *P*_FDR_ = .362; wave-4, β = 0.079, SE = 0.099, *P*_FDR_ = .424; dashed lines in Figure 3B), but remained significant in all other subgroups (*P*_FDR_ < .05; Supplementary Table 8.2).

**Figure 3.**
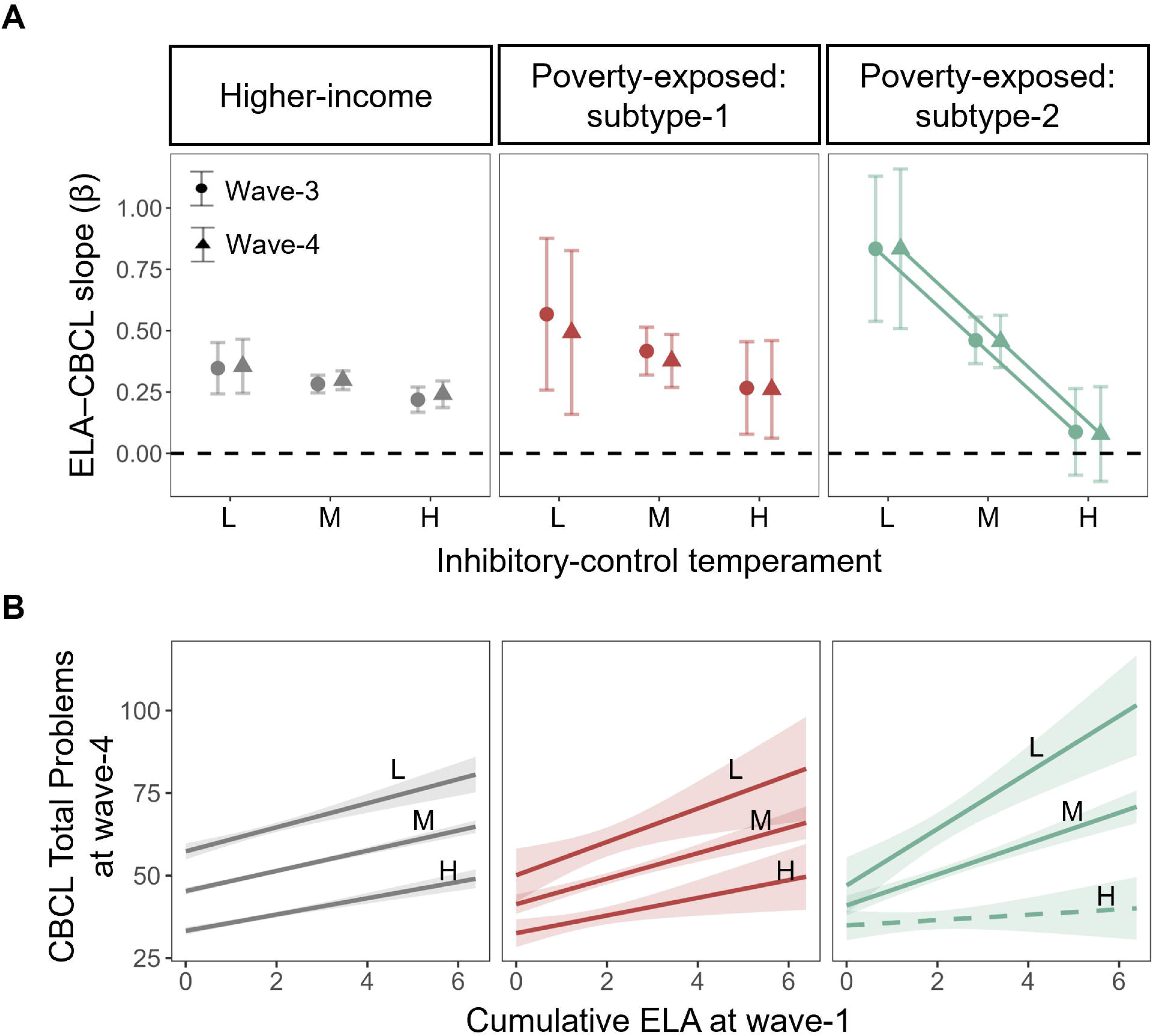
Temperamental Inhibitory Control and Neurofunctional Subtype Jointly Stratify Adversity–Psychopathology Coupling. **A,** Model-derived ELA–CBCL slopes (β) at waves-3 and −4 across levels of temperamental inhibitory control, shown separately for higher-income youth and the 2 poverty-exposed subtypes. Temperament levels are displayed as low (L), medium (M), and high (H), corresponding to EATQ-R inhibitory-control subscale values of 1, 3, and 5 (range, 1–5). Circles indicate wave-3 and triangles indicate wave-4. Points indicate estimated marginal ELA–CBCL slopes; error bars represent 95% CIs. The dashed horizontal line denotes β = 0. **B,** Wave-4 model-implied associations between baseline cumulative ELA burden (wave-1) and CBCL Total Problems at wave-4, shown separately by group and temperament level (L/M/H defined as EATQ-R values 1/3/5). Lines depict estimated marginal associations; shaded bands represent 95% CIs. The dashed regression line indicates a non-significant association between cumulative ELA at wave-1 and CBCL Total Problems at wave-4. Abbreviations: CBCL, Child Behavior Checklist; ELA, early-life adversity; EATQ-R, Early Adolescent Temperament Questionnaire–Revised; CI, confidence interval. Alt text: Plots show inhibitory-control temperament and fMRI subtype jointly moderate ELA–CBCL slopes; buffering appears in subtype-2 with high control.

### Sensitivity Analyses

In sensitivity analyses, the primary pattern of group differences in cumulative-ELA–related vulnerability (the slope of CBCL Total Problems on cumulative ELA) remained consistent across three alternative specifications. Findings were materially unchanged (1) when cumulative ELA was defined using all 14 adversity indicators rather than the primary 8 indicators showing poverty-amplified associations (Supplementary Results 3.1), (2) when analyses were restricted to complete cases without imputed ELA values (Supplementary Results 3.2), (3) and when only one child per family was retained (Supplementary Results 3.3). Across these analyses, poverty continued to show steeper adversity–psychopathology coupling than higher-income peers. Subtype-1 remained more vulnerable than the higher-income group, whereas subtype-2 retained an attenuated-vulnerability profile, and the strongest temperamental buffering persisted within subtype-2.

### Molecular Correspondence

Exploratory multiscale molecular mapping contextualized subtype-specific stop-signal activation differences (Figure 4A). Using spin-based inference (10,000 rotations) to account for spatial autocorrelation, the poverty subtype-1 (high-activation) versus higher-income β difference map showed positive correspondence with PET-derived monoaminergic receptor topographies, including serotonergic 5-HT1A (*r* = .202; *P*_spin_ = .048; Figure 4G) and dopaminergic D2 distributions (D_21_: *r* = .177, *P*_spin_ = .050; D_23_: *r* = .209, *P*_spin_ = .042). In contrast, poverty subtype-2 (low-activation) showed no significant receptor/transporter correspondence (Figure 4F; Supplementary Table 9). Complementary transcriptomic mapping linked subtype β difference maps to Allen Human Brain Atlas cortical expression profiles via partial least squares regression: PLS1 exhibited significant covariance for subtype-2 (variance explained = 38.6%; *P*_spin_ = .024; Figure 4B and 4D) but not subtype-1 (30.7%; Pspin = .233), indicating a subtype-specific transcriptomic correspondence. Genes contributing to the subtype-2 pattern were enriched for glial and precursor cell classes (astrocytes, oligodendrocytes, microglia, and OPCs; all P < .0001; Figure 4E) and underrepresented for neuronal classes (excitatory and inhibitory; P < .0001), providing preliminary biological context for the subtype that showed transcriptomic but not receptor/transporter correspondence. These results are detailed in Supplementary Results 4.

**Figure 4.**
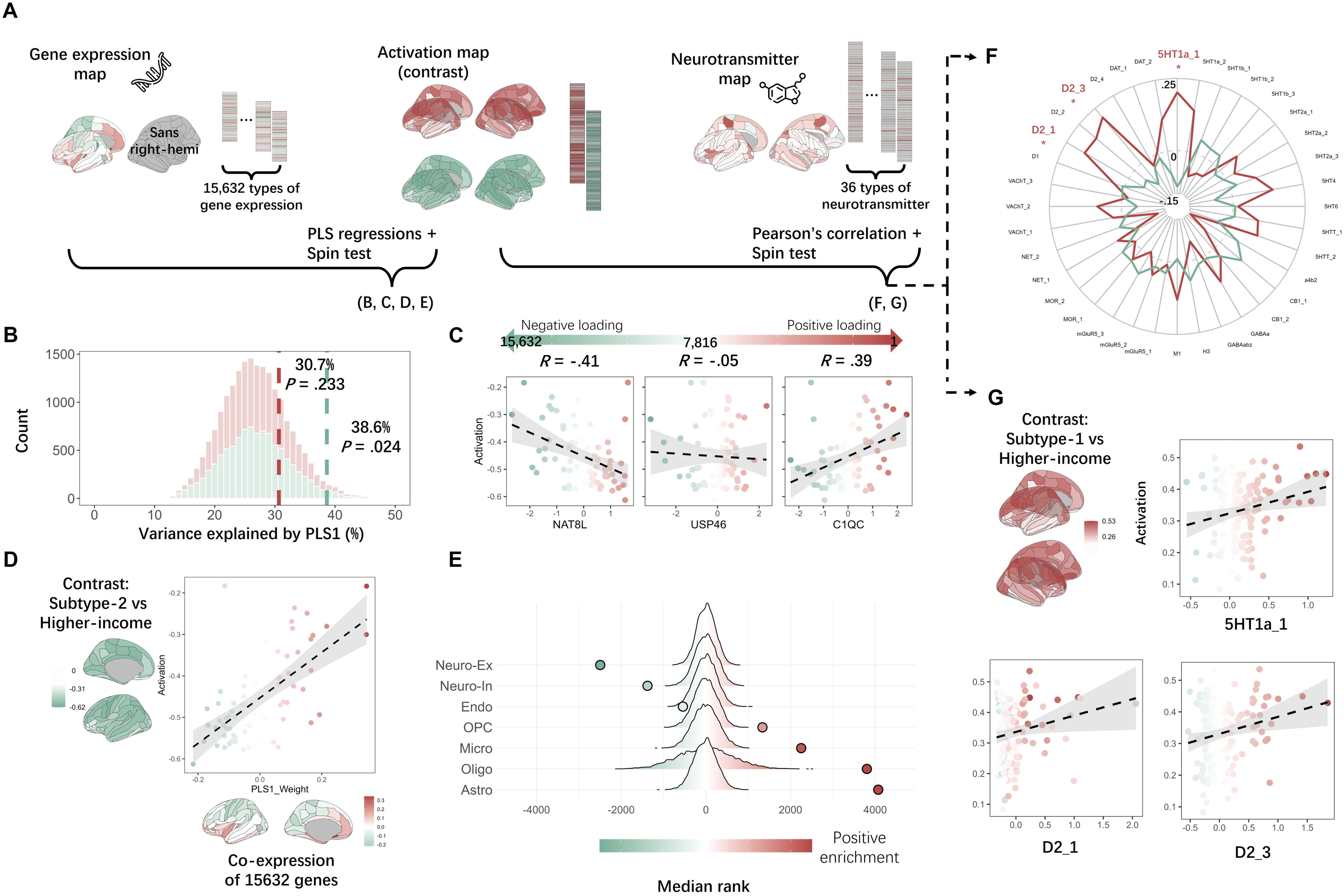
Multiscale Molecular Correspondence of Neurofunctional Subtype β Difference Maps. **A,** Overview of exploratory multiscale mapping analyses linking subtype-specific cortical subtype–higher-income β difference maps (standardized coefficients) to AHBA transcriptomic data and PET-derived neurotransmitter receptor/transporter distributions. Transcriptomic analyses used 15,632 genes across 72 left-hemisphere Destrieux regions (right-hemisphere coverage is incomplete in AHBA). PET analyses used 36 cortical receptor/transporter maps. Statistical inference used spin-based permutation testing (10,000 rotations) to account for spatial autocorrelation. **B,** Spin-based null distributions for the variance explained by PLS1 in transcriptomic analyses. Vertical dashed lines indicate observed variance explained for poverty subtype-1 (30.7%; *P*_spin_ = .233) and poverty subtype-2 (38.6%; *P*_spin_ = .024). **C,** Examples of genes spanning the PLS1 weight distribution (negative to positive loadings), illustrating regional associations between gene expression and subtype-specific β differences across left-hemisphere regions. **D,** Spatial correspondence between the poverty subtype-2 subtype–higher-income β difference map and PLS1 weights across left-hemisphere regions, with accompanying cortical projection of PLS1 weights. **E,** Cell-type enrichment results for genes ranked by PLS1 weights in poverty subtype-2, shown as median-rank summaries across 7 broad cell classes. **F,** Summary of receptor/transporter correspondence for poverty subtype-1 and poverty subtype-2, shown as Pearson correlations between each subtype-specific β difference map and each PET receptor/transporter distribution (spin-test inference), with illustrative significant correspondences for poverty subtype-1 (5-HT1A: *r* = .202, *P*_spin_ = .048; D2 distributions: D_21_ *r* = .177, *P*_spin_ = .050; D_23_ *r* = .209, *P*_spin_ = .042). Abbreviations: AHBA, Allen Human Brain Atlas; PET, positron emission tomography; PLS, partial least squares. Alt text: Schematic and plots show exploratory mapping links subtype activation-difference maps to PET receptor distributions and AHBA gene expression; subtype-1 aligns with monoaminergic receptors, whereas subtype-2 shows a transcriptomic signature with cell-type enrichment.

## Discussion

Childhood poverty is widely recognized as a complex high-risk context for mental health. Here, in a US cohort followed from late childhood into adolescence, childhood poverty amplified adversity-linked vulnerability. Critically, within poverty-exposed youth, a discriminative clustering approach identified two neurofunctional subtypes that were broadly similar on key non-neural characteristics at baseline, yet diverged into distinct vulnerability and resilience profiles over follow-up. Temperamental inhibitory control further refined this divergence, with the most buffered pattern observed among youth with high temperament and the lower-activation profile. Exploratory molecular mapping provided biological context for these subtype-defined profiles.

A steeper adversity–psychopathology coupling under childhood poverty fits influential stress frameworks. Persistent strain in the context of poverty can tax regulatory systems, increasing the likelihood that subsequent stressors translate into psychopathology.^2^ In parallel, economic-hardship models specify family pathways through which financial pressure disrupts caregiving processes and buffering resources that support children’s adjustment and self-regulation.^23^ Our findings translate these frameworks into a measurable developmental index by estimating vulnerability as the slope linking adversity accumulation to symptom growth. This emphasis matters in high-risk settings where symptom levels may be influenced by prior experiences and context.^24^ Mean-level differences can then be a less specific readout of system functioning, particularly when cross-sectional analyses blend prior accumulation with measurement and reporting context.^25^ By focusing on the longitudinal slope, we more directly quantify adversity-linked vulnerability to accumulating adversity, consistent with stress-sensitization accounts in which early adversity amplifies later symptom reactivity.^26^

Within poverty-exposed youth, multivariate clustering of distributed stop-signal task (SST)–evoked activation delineated two neurofunctional profiles that were broadly similar on key non-neural characteristics, yet diverged in adversity–psychopathology coupling over time. This kind of person-level, brain-based stratification is increasingly recommended to address heterogeneity that is not well resolved by group averages, especially when mechanisms are expressed as distributed patterns rather than isolated loci.^27^ Consistent with that rationale, recent work in adolescent depression has used neurobiological subtypes to differentiate clinically meaningful profiles, underscoring the value of brain-based stratification in youth psychopathology.^28^ We extend this stratification to an exposure-defined setting where neurodevelopmental and mental-health outcomes can diverge substantially across individuals.^3^ In doing so, we introduce a mechanism-grounded, within-poverty stratification approach that goes beyond group averages and helps identify who is more likely to show escalating problems as adversities accumulate, thereby informing precision-oriented prevention and evaluation.

These results also sharpen the interpretation of resilience-like functioning in children growing up in poverty. Resilience is defined as sustained or recovered mental health despite chronic contextual risk,^29^ and longitudinal work shows that many exposed individuals follow stable-adaptive or recovery trajectories rather than uniform symptom elevation.^30^ By estimating adversity–psychopathology slope, we isolate a dynamic signature of vulnerability that also clarifies how resilience-like functioning can emerge as attenuated adversity-to-psychopathology translation over time.^31^ We identified a poverty-exposed subgroup in which higher adversity burden was followed by markedly less symptom growth, indicating a buffered, resilience-like profile across follow-up. Inhibitory control is a plausible leverage point because it governs prepotent responding and is consistently implicated in pathways linking early adversity and socioeconomic strain to later mental-health risk.^32^ It may therefore represent a candidate process underlying buffered vulnerability and resilience-like functioning under childhood poverty.

Childhood poverty may recalibrate distributed control systems that support goal-directed regulation and inhibition.^33^ Chronic psychosocial strain has been associated with altered computations in control networks, including prefrontal processes supporting inhibitory control.^34^ Within mechanistic accounts of cognitive control, stronger task-evoked recruitment is not necessarily advantageous and may index compensatory allocation under higher demands.^16^ Consistent with this view, the higher-activation subtype’s steeper adversity–psychopathology slope may reflect heightened vulnerability under rising regulatory demands, whereas the lower-activation subtype may reflect a more sustainable, buffered configuration of control deployment. A common interpretation is that sustained pressure compromises control efficiency and increases the control costs required to maintain goal-directed performance under demand.^35^ Critically, however, this pattern may not represent a simple deficit, but rather a contextual adaptation. Such high-effort recruitment may reflect system-level recalibration to sustain functioning under environmental strain, but it may also carry allostatic costs that undermine the maintenance of resilience-like functioning over time.^9^

Temperamental inhibitory control complemented the neurofunctional profiles, with the most buffered profile emerging selectively when higher temperamental control co-occurred with the lower-activation neurofunctional profile. This pattern indicates that behavioral self-control and control-network recruitment jointly differentiate vulnerability and resilience profiles within poverty, rather than providing redundant markers of the same construct. The moderation implies that the protective value of everyday self-control is not uniform within poverty-exposed youth; it is strongest when lower-cost neurofunctional engagement accompanies higher temperamental control.^36^ Prospective evidence indicates that earlier self-control predicts a wide range of later-life outcomes, including health and socioeconomic functioning, supporting an everyday marker of regulatory capacity as informative for long-horizon stratification of vulnerability and resilience.^37^ At the same time, training among lower-socioeconomic-status preschoolers has been shown to improve neurocognitive function and behavior, supporting the premise that regulatory systems can be strengthened in constrained contexts.^38^ Thus, the subtype-by-temperament interaction offers a practical bridge between scalable behavioral screening and neurobiological characterization.^29^

Exploratory molecular mapping provided additional biological context for these subtype patterns. The higher-vulnerability profile showed spatial correspondence with serotonergic and dopaminergic receptor systems, which are broadly implicated in stress responsivity and mood-related risk.^39^ The buffered, resilience-like profile showed transcriptomic correspondence that was more consistent with glial and precursor cell signatures, aligning with evidence that glia–neuron interactions contribute to stress-related plasticity.^40^ Although spatial correspondence does not establish mechanism, these maps help prioritize candidate molecular systems for follow-up studies that can more directly test causal pathways linking poverty-related strain, control-network configuration, and symptom trajectories.

We acknowledge several limitations. First, mental-health outcomes were assessed via parent-reported CBCL. Although the CBCL shows strong reliability and validity,^22^ parent ratings are subject to systematic reporting biases, including those linked to family income.^1^ Future work incorporating youth self-report, teacher report, and clinical interviews will be important for testing whether these findings generalize across informants and assessment modalities. Second, our analyses focused on mental-health outcomes, while prior research shows that psychological success in poverty contexts can sometimes come at the expense of physical health.^4,9^ Long-term follow-ups measuring physical-health indicators are necessary to determine whether this buffered, resilience-like profile is truly health-promoting in the long run, or instead involves trade-offs in other domains. Although ABCD is community based, the analytic sample is not fully representative and is majority White; findings may not generalize to groups exposed to different structural inequities. Race and ethnicity were treated as social constructs; future work should test whether structural racism and context modify adversity exposure and adversity-linked vulnerability.

In conclusion, childhood poverty amplified adversity-linked vulnerability over development, and multilevel inhibitory-control profiles stratified vulnerability and resilience within poverty-exposed youth. By framing vulnerability as a longitudinal slope and showing that this slope is prospectively separable by neurobehavioral inhibitory-control profiles, these findings provide a developmentally grounded framework for within-poverty stratification. These findings may help identify which youth are most likely to translate adversity burden into later symptom burden, even within similarly disadvantaged contexts. They also highlight inhibitory control as a candidate protective process for prevention, suggesting that assessments of everyday self-regulation, potentially combined with neurocognitive indicators, may improve developmental risk stratification. More broadly, the results support prevention strategies that strengthen self-regulatory capacity and supportive caregiving environments in families and schools, rather than treating poverty exposure itself as a uniform proxy for poor outcome.

## Data Availability

Data used in this study are available from the NIMH Data Archive (ABCD Study) to qualified investigators upon approval of a Data Use Certification. Analytic code will be made available from the corresponding author upon reasonable request / in a public repository at acceptance.

