## Supplementary material for "Neurobehavioral Profiles of Inhibitory-Control Stratify Vulnerability and Resilience under Childhood Poverty"

**Supplementary Materials**

**Methods 1.** Sample selection and analytic cohorts

**Methods 2.** Poverty exposure and early-life adversity

**Methods 3.** Other measurements

**Methods 4.** Neuroimaging processing and quality control

**Methods 5.** Neurofunctional subtyping with HYDRA

**Methods 6.** Statistical modeling of adversity–problem coupling

**Methods 7.** Sensitivity and robustness analyses

**Methods 8.** Exploratory multiscale molecular mapping

**Methods 9.** Neurotransmitter receptor/transporter correspondence

**Methods 10.** Transcriptomic correspondence

**Results 1.** Structural MRI differences between subtypes

**Results 2.** Stop-signal task performance and behavioral moderation checks

**Results 3.** Sensitivity analyses

**Results 3.1** Sensitivity analysis 1: cumulative ELA defined using all 14 ELAs

**Results 3.2** Sensitivity analysis 2: cumulative ELA without imputed missing values (complete-case)

**Results 3.3** Sensitivity analysis 3: cumulative ELA after random sibling exclusion

**Results 4.** Multiscale molecular correspondence

**Table 1.** Associations between each early-life adversity (ELA) indicator and CBCL total problems (ELA–CBCL coupling) by poverty and interaction tests

**Table 2.** Standardized associations between cumulative ELA and CBCL total problems (cumulative ELA–CBCL coupling) across waves, stratified by poverty status (before fMRI QC) and neurofunctional subtype (after fMRI QC)

**Table 3.** Differences in cumulative ELA–CBCL coupling between the poverty-exposed and higher-income groups across waves

**Table 4.** Regional differences in stop-signal task activation between poverty subtypes and the higher-income group

**Table 5.** Participant characteristics across the higher-income group and poverty subtypes in the task-fMRI analytic sample

**Table 6.** Differences in cumulative ELA–CBCL coupling between poverty neurofunctional subtype-1 and subtype-2 across waves

**Table 7.** Regions showing significant (uncorrected) interactions with cumulative ELA and poverty status in predicting CBCL total problems

**Table 8.** Moderation of cumulative-ELA-CBCL associations by inhibitory-control temperament and poverty/subtype status

**Table 9.** Spatial correlations between subtype activation patterns and neurotransmitter receptor/transporter distributions

**Methods 1. Sample selection and analytic cohorts**

Analyses used the ABCD baseline cohort with follow-up assessments through adolescence. The primary “full cohort” analyses used all participants with complete demographic data and at least one assessment of CBCL total problems. Poverty status was defined at baseline using the federal poverty threshold based on household income and family size, yielding a poverty-exposed group and a higher-income comparison group. For analyses involving task-fMRI activation, participants additionally required usable stop-signal task fMRI data after standard quality-control procedures.

**Methods 2. Poverty exposure and early-life adversity**

Poverty exposure was defined at baseline using the federal poverty threshold derived from household income and family size (income-to-needs ratio < 1.0). Early-life adversity (ELA) was operationalized using 14 harmonized indicators spanning prenatal, child health, family, school, and neighborhood domains. We evaluated both ELA-specific vulnerability (poverty moderation of each ELA–CBCL slope) and cumulative ELA vulnerability, using a primary cumulative index based on ELAs whose effects were amplified under poverty and sensitivity indices based on all 14 indicators.

**Poverty exposure.** Household income and family size were used to determine whether the participant’s household fell below the federal poverty threshold (income-to-needs ratio) at baseline. Poverty exposure was defined by an income-to-needs ratio below 1.0 at baseline (i.e., household income relative to the federal poverty threshold given family size), treated as a binary indicator in the primary analyses.

**Early-life adversity (ELA).** We operationalized ELA using 14 standardized indicators spanning multiple domains:

**(1) Surgery (categorical).** Caregivers reported whether the child had any surgery (0 = no, 1 = yes).

**(2) Prenatal substance exposure (continuous).** Prenatal exposure was operationalized as the sum of six binary exposures that were each independently associated with small but significant increases in CBCL total scores: unplanned pregnancy; maternal alcohol use early in pregnancy; maternal marijuana use early in pregnancy; maternal tobacco use early in pregnancy; pregnancy complications; birth complications.^1^ The number of exposures (0-6) was summed and rescaled linearly to a 0-1 range.

**(3) Experience of trauma (categorical).** Caregivers reported whether the child had experienced any potentially traumatic event (including serious accident, serious injury/medical emergency, exposure to a disaster, witnessing violence, being a victim of violence, and other traumatic events). Exposure was coded as 1 if any event was endorsed and 0 otherwise.

**(4) Parental history of substance abuse (categorical).** Caregiving reported whether either of the child’s parents had alcohol problems or drug-use problems. If at least one parent had either an alcohol or a drug-use problem, the child was coded as 1; otherwise, exposure was coded as 0.

**(5) Parental separation (categorical).** Parents reported their current marital status (1 = Married, 2 = Widowed, 3 = Divorced, 4 = Separated, 5 = Never married, 6 = Living with a partner). Children whose responding caregiver was neither married (1) nor living with a partner (6) were coded as 1; all others were coded as 0.

**(6) Area deprivation index (continuous).** Neighborhood deprivation was indexed using a geocoded Area Deprivation Index (ADI) linked to participants’ residential location; ADI values were rescaled to 0–1 with higher values indicating greater neighborhood deprivation.

**(7) Neighborhood unsafety (continuous).** Caregivers reported neighborhood safety using the ABCD Neighborhood Safety/Crime Survey; scores were rescaled to 0–1 with higher values indicating greater neighborhood unsafety.

**(8) Sleep problems (continuous).** Caregivers reported sleep difficulties using the ABCD Sleep Disturbance Scale for Children; scores were rescaled to 0–1 with higher values indicating greater sleep problems.

**(9) Adverse school environment (continuous).** Youth reported perceived school environment (e.g., safety, support, and climate); scores were rescaled to 0–1 with higher values indicating a more adverse school environment.

**(10) Peer bullying (categorical).** Parents reported whether the child had problems with bullying at school or in the neighborhood (“Does your child have any problems with bullying at school or in your neighborhood?”; 1 = Yes, 0 = No). A response of 1 was coded as exposure.

**(11) Family conflict (continuous).** Caregivers completed the Family Environment Scale (FES) conflict subscale; scores were rescaled to 0–1 with higher values indicating greater family conflict.

**(12) Parent psychopathology (continuous).** Parental psychopathology was measured using the Adult Self Report (ASR) total-problems T-score for the primary caregiver. T-scores were provided by ABCD team based on national norms and then rescaled to a 0-1 range, with higher values indexing higher self-reported psychopathology.

**(13) Parental neglect (continuous).** Youth completed a Parental Monitoring Survey assessing perceived neglect/low monitoring; scores were rescaled to 0–1 with higher values indicating greater neglect.

**(14) Parental coldness (continuous).** Youth completed a Parental Warmth Survey assessing perceived parental coldness/low warmth; scores were rescaled to 0–1 with higher values indicating greater parental coldness.

Binary indicators were coded as 0/1 for absence/presence. Continuous indicators were linearly rescaled to a 0–1 range (higher values indicate greater adversity burden) to facilitate aggregation across domains.

**Missing ELA values.** For the primary cumulative indices, missing values on individual ELA indicators were imputed using the sample mean for that indicator prior to summation; we then conducted complete-case sensitivity analyses without imputation (see Supplementary Methods 7 and Supplementary Results 3).

**Primary cumulative ELA index (“8-ELA index”).** In the primary framework, cumulative ELA burden was operationalized using the subset of 8 ELA indicators that exhibited significantly stronger prospective associations with CBCL total problems in the poverty-exposed group in ELA-specific interaction models (7 of these 8 interactions survived FDR correction, Supplementary Table 1). Each indicator was rescaled to range from 0 to 1 and summed, yielding an index in which higher values reflect greater cumulative burden within this poverty-sensitive subset. Sensitivity analyses additionally tested (i) an alternative cumulative ELA index computed as the sum of all 14 indicators, (ii) models excluding participants with any imputed ELA indicator(s), and (iii) models excluding one randomly selected sibling from families contributing multiple children (Supplementary Results 3).

**Methods 3. Other measurements**

**Behavioral problems.** Children’s mental-health outcomes were assessed at baseline and at three subsequent annual follow-ups using the parent-report Child Behavior Checklist.^2^ The CBCL includes 113 items assessing behavioral and emotional problems over the past six months, rated on a 3-point scale (0 = Not True, 1 = Somewhat or Sometimes True, 2 = Very True or Often True). Item scores were summed to form syndrome and broadband scales, and converted to age- and sex-normed T-scores. We used the CBCL total problems T-score as the primary outcome indexing overall behavioral problems in all analyses of ELA-related vulnerability. In the primary framework, ELA-related vulnerability was operationalized as the standardized regression coefficient linking cumulative ELA to CBCL total problems.

**Temperamental inhibitory control.** Temperamental inhibitory control was assessed using the 5-item Inhibitory Control subscale from the Early Adolescent Temperament Questionnaire–Revised (EATQ-R). Caregivers completed the EATQ-R at wave-3. Items were averaged to create a scale score, with higher values indicating greater temperamental inhibitory control. For visualization and simple-slope probing, conditional effects were evaluated at scale values of 1, 3, and 5 on the 1–5 EATQ-R inhibitory-control subscale, corresponding to low, medium, and high temperament.

**Cognitive assessments.** Cognitive function was assessed using the NIH Toolbox Cognition Battery. We examined both fluid cognition and crystallized cognition composites, as well as the seven task measures that contribute to these composites: List Sorting Working Memory, Flanker Inhibitory Control and Attention, Dimensional Change Card Sort, Pattern Comparison Processing Speed, Picture Sequence Memory, Picture Vocabulary, and Oral Reading Recognition. For analyses, we used age-corrected standard scores to reduce confounding by age differences across participants. Measures and scoring conventions followed ABCD study documentation for the corresponding release (see Supplementary Table 7).

**Stop-signal task fMRI activation.** Inhibitory-control–related brain function was indexed using task-evoked activation during the stop-signal task. Activation features were summarized as ROI-level values derived from an established cortical atlas and subcortical segmentation, yielding a multivariate activation vector for each participant. The ROI approach was chosen to capture distributed activation patterns.

**Methods 4. Neuroimaging processing and quality control**

All MRI data in the ABCD Study were acquired on harmonized 3T protocols across 21 sites and centrally preprocessed and quality-controlled by the ABCD imaging team.^3,4^ Structural T1-weighted images were corrected for gradient nonlinearity and intensity inhomogeneity, registered to standard space, and processed with FreeSurfer to derive ROI-level measures. Cortical regions were defined using the Destrieux atlas (148 ROIs), and subcortical structures using the ASEG atlas (30 ROIs), yielding cortical thickness and regional volume estimates per ROI. For structural analyses in the present study, we required that T1-weighted data meet the ABCD recommended inclusion criterion.

Stop-signal task (SST) fMRI data were preprocessed using the ABCD task-fMRI pipeline.^4^ Preprocessing included removal of initial volumes, correction for gradient nonlinearity and B0 field distortions using field maps, rigid-body motion correction (realignment to a reference image), registration of each run’s mean BOLD image to the participant’s T1-weighted image, transformation to a common template space, and projection to the cortical surface. Temporal filtering and intensity normalization were applied to reduce low-frequency drift and inter-run intensity differences. Head motion was quantified using framewise displacement (FD) and related metrics within the ABCD pipeline. For primary SST analyses, we followed ABCD recommendations and included only runs meeting the standardized inclusion criterion.

First-level SST task-fMRI analyses were implemented as general linear models (GLMs) estimated on the preprocessed BOLD time series (Hagler et al., 2019). Regressors were defined for trial types (e.g., correct go, correct stop, and error trials) and convolved with a canonical hemodynamic response function. Models included nuisance regressors for head-motion parameters and low-frequency trends, and high-motion volumes were censored within the design matrix. The ABCD pipeline produced participant-level contrast maps. Consistent with prior SST work in ABCD,^5–7^ we focused on the correct stop vs correct go contrast as the primary index of inhibitory-control-related activation.

For each participant, we extracted ROI-level beta estimates for the correct stop vs correct go contrast by averaging contrast values within each Destrieux cortical parcel and ASEG subcortical structure, yielding a 178-dimensional activation vector (148 cortical + 30 subcortical ROIs). These multivariate activation features served as inputs for HYDRA-based subtyping and for mixed-effects models of cumulative-ELA-related vulnerability. In models with ROI-level activation as the dependent variable, we additionally adjusted for run-level mean FD; in models of cortical thickness, we further adjusted for mean cortical thickness across ROIs to account for global differences.

**Methods 5. Neurofunctional subtyping with HYDRA**

We used HYDRA (Heterogeneity Through Discriminative Analysis),^8^ a semi-supervised machine-learning algorithm that combines binary classification with subtype clustering. HYDRA learns a convex polytope defined by multiple linear hyperplanes that jointly separate a target (“clinical”) group from a reference group; each target-group participant is assigned to the hyperplane on which they are best discriminated, thereby defining subtypes.

In our application, children from families below the federal poverty threshold were treated as the target group and those at or above the threshold as the reference group. Input features were task-evoked activation values (beta weights) for the stop-signal task inhibitory-control contrast (correct stop vs correct go) across all cortical and subcortical ROIs (178 features). Prior to model fitting, we regressed out age, birth sex, head motion (mean framewise displacement), race andethnicity, and study site from the activation features.

We evaluated HYDRA models specifying 2–5 clusters using 10-fold cross-validation. For each solution, we computed the adjusted Rand index (ARI) to quantify clustering stability. The 2-cluster model yielded the highest ARI (0.505), indicating that two neurofunctional subtypes provided the most stable characterization of inhibitory-control activation patterns in the poverty-exposed group.

To assess statistical significance, we performed a permutation test in which poverty-exposure labels were randomly shuffled to generate null models. We generated 100 null-label datasets and refit the 2-cluster HYDRA model to each. The ARI from the empirical model exceeded the null distribution (*P* < .05), supporting the robustness of the 2-subtype solution.

**Methods 6. Statistical modeling of adversity–problem coupling**

We estimated adversity–problem coupling (ie, the ELA–CBCL slope) in 2 complementary frameworks: (1) a cross-sectional baseline analysis (wave 1) and (2) longitudinal analyses that jointly modeled repeated CBCL assessments across waves 1–4.

In baseline models, we regressed baseline CBCL Total Problems on baseline cumulative ELA, the moderator of interest (poverty status or HYDRA subtype), and their interaction, adjusting for baseline age, sex at birth, race and ethnicity, and continuous income-to-needs (log-transformed), with a random intercept for recruitment site and family (family nested within site). Baseline ELA–CBCL slopes within each poverty group (or subtype) were obtained as model-implied simple slopes (estimated marginal trends) from the ELA × moderator interaction.

In longitudinal models, we fit a single linear mixed-effects model to all available CBCL observations across waves 1–4, including fixed effects for time (wave), baseline poverty status (or subtype), baseline cumulative ELA, and their interactions (group × wave × ELA), with random intercepts for recruitment site and family (family nested within site) and a participant-level random intercept to account for repeated measurements. We additionally adjusted for visit age and continuous income-to-needs (log-transformed). Wave-specific ELA–CBCL slopes were derived from the longitudinal model as model-implied simple slopes at each wave within each group (i.e., conditional ELA slopes stratified by wave). For both baseline and longitudinal models, inference focused on interaction terms and on contrasts of model-implied ELA slopes between groups (or between subtypes) as appropriate.

**Methods 7. Sensitivity and robustness analyses**

We conducted three prespecified sensitivity analyses to evaluate whether the primary conclusions were robust to (i) how cumulative ELA was constructed, (ii) the handling of missing ELA values, and (iii) residual family-level clustering due to siblings/twins. All sensitivity models followed the same statistical specifications as the primary analyses (Supplementary Methods 6), including identical covariates and interaction tests, and were evaluated in the same cross-sectional (baseline) and wave-specific follow-up frameworks when applicable.

**Sensitivity analysis 1: cumulative ELA defined using all 14 ELAs**

In the primary analyses, cumulative ELA was defined using the eight ELA domains that showed significant poverty × ELA interactions on CBCL total problems in cross-sectional models, emphasizing ELA types whose effects were most clearly moderated by poverty status. To test whether results depended on this selection, we recomputed cumulative ELA as the sum of all 14 ELA indicators and repeated the key tests: (i) poverty × cumulative ELA → CBCL total problems, (ii) subtype × cumulative ELA → CBCL total problems (benchmarking each poverty subtype against the higher-income comparison group), and (iii) subtype × inhibitory-control temperament × cumulative ELA → CBCL total problems at follow-ups.

**Sensitivity analysis 2: complete-case cumulative ELA without mean imputation**

In the primary analyses, missing values for individual ELAs were mean-imputed before computing cumulative ELA. To test whether imputation influenced results, we recomputed cumulative ELA from the eight key ELA indicators using only participants with complete data on all eight indicators (no imputation). Baseline missingness among the 10,112 children in the main sample was: parental substance-use problems, 490 (4.8%); experience of trauma, 266 (2.6%); experience of bullying, 2 (0.02%); experience of surgery, 1 (0.01%); parental psychopathology, 54 (0.53%); parental coldness, 0; sleep problems, 202 (2.0%); and prenatal exposure, 1,432 (14.2%). After excluding participants with missing data on any of these eight indicators, 8,094 children remained for complete-case analyses. We then repeated: (i) poverty × cumulative ELA → CBCL total problems, (ii) subtype × cumulative ELA → CBCL total problems, and (iii) subtype × inhibitory-control temperament × cumulative ELA → CBCL total problems.

**Sensitivity analysis 3: random sibling exclusion**

The ABCD Study oversampled twins and siblings.^9^ In our main sample (N = 10,112), 1,700 children (16.8%) had at least one sibling also enrolled. Therefore, we conducted an additional robustness test that removed sibling clusters by randomly retaining only one child per family. After random sibling exclusion, 8,414 children remained at baseline. We repeated the same key tests as above: (i) poverty × cumulative ELA → CBCL total problems, (ii) subtype × cumulative ELA → CBCL total problems, and (iii) subtype × inhibitory-control temperament × cumulative ELA → CBCL total problems.

**Methods 8. Exploratory multiscale molecular mapping**

We conducted prespecified, hypothesis-generating multiscale mapping analyses to contextualize subtype-specific stop-signal activation patterns at the molecular level (workflow shown in Figure 4A). For each neurofunctional subtype, we first derived a subtype-specific cortical subtype–higher-income difference map defined by standardized coefficients (β) estimated from models comparing that subtype with the higher-income comparison group. These subtype-specific β maps were then projected onto the cortical surface for molecular correspondence analyses. We then tested spatial correspondence between these β difference maps and (1) PET-derived neurotransmitter receptor/transporter distributions and (2) cortical gene-expression profiles from the Allen Human Brain Atlas (AHBA). All molecular analyses were restricted to the cortical surface and used spin-based permutation procedures (10,000 rotations) to preserve spatial autocorrelation. Because these molecular atlases are derived from adult samples and provide spatial correspondence rather than individual-level inference, all results were interpreted as exploratory and hypothesis-generating.

**Methods 9. Neurotransmitter receptor/transporter correspondence**

To test whether subtype-specific activation patterns were spatially aligned with particular neurotransmitter systems, we related subtype subtype–higher-income β difference maps (standardized coefficients) to PET-derived cortical distributions of neurotransmitter receptors and transporters. We used PET-derived distribution maps aggregated from 36 receptor/transporter maps covering 19 receptor and transporter types in approximately 1,200 healthy adults.^10^ These maps were parcellated into Destrieux cortical ROIs, excluding subcortical structures.

For each subtype, we derived a subtype-specific subtype–higher-income β difference map (standardized coefficients) and computed Pearson correlation coefficients between this β map and each receptor/transporter distribution. Statistical significance was evaluated using 10,000 spin permutations that preserve the spatial autocorrelation structure of the cortical surface, implemented via the gen_spinsamples function in the netneurotools package.^10^ Corresponding correlation coefficients and spin-test p-values are reported in Supplementary Table 9, and illustrative results are shown in Figure 4F and 4G.

**Methods 10. Transcriptomic correspondence**

To link subtype-specific activation patterns to transcriptional profiles, we used cortical gene-expression data from the Allen Human Brain Atlas (AHBA; https://human.brain-map.org/), which provides high-resolution spatial expression data for thousands of genes across six postmortem adult brains (4 male, 2 female; mean age ≈ 45 years). AHBA data were preprocessed using the abagen toolbox (default settings; https://github.com/rmarkello/abagen) and parcellated into Destrieux cortical ROIs. Because right-hemisphere coverage is incomplete in AHBA, transcriptomic analyses were restricted to the 72 left-hemisphere ROIs with available expression data, yielding a matrix of 15,632 genes across 72 regions (overview in Figure 4A).

For each subtype, partial least squares (PLS) regression related the gene-expression matrix to the subtype-specific subtype–higher-income β difference vector (standardized coefficients) across these 72 regions (genes as predictors; regional β differences as the response). We extracted the first PLS component (PLS1) as the major axis of shared variance between activation and expression. To assess significance, we generated a null distribution for the variance explained by PLS1 using 10,000 spin permutations that preserved spatial autocorrelation (null distributions shown in Figure 4B). PLS1 was considered significantly associated with the β difference map if the observed variance explained exceeded the 95th percentile of the spin-based null distribution. In the present study, this criterion was met for poverty subtype-2 but not for poverty subtype-1 (Figure 4B).

For the subtype with a significant PLS1 association (poverty subtype-2), we conducted cell-type enrichment analysis to test whether genes with stronger PLS1 weights were preferentially expressed in specific cell classes. Using a meta-analysis of cell-type-specific transcriptional signatures,^11^ genes were assigned to one of seven broad cell types: excitatory neurons, inhibitory neurons, endothelial cells, astrocytes, microglia, oligodendrocytes, and oligodendroglial precursor cells (OPCs). We ranked genes by their PLS1 weights and computed the median rank of genes associated with each cell type. Observed median ranks were compared with null distributions obtained via spin permutations; a cell type was considered significantly enriched (positively or negatively) if its median rank lay outside the central 95% of the null distribution (cell-type results shown in Figure 4E). Illustrative gene-level associations and spatial correspondence patterns are shown in Figure 4C and 4D. All transcriptomic results were interpreted as exploratory because the atlases are derived from adult samples and provide spatial correspondence rather than individual-level or causal inference.

**Results 1. Structural MRI differences between subtypes**

At the level of brain structure, no cortical-thickness or volumetric differences survived FDR correction. At a more liberal uncorrected threshold (*P* < .01), two focal cortical-thickness differences emerged: compared with children with poverty subtype-1 (higher-activation profile), those with poverty subtype-2 (lower-activation profile) showed thinner cortex in the left middle frontal gyrus (mean ± SE = 2.956 ± 0.008 vs. 2.974 ± 0.008 mm; Δ = 0.017 mm; t = 2.78, *P* = .005) and the right angular gyrus (mean ± SE = 2.979 ± 0.009 vs. 3.008 ± 0.009 mm; Δ = 0.029 mm; t = 3.80, *P* = .0001). No volumetric differences were observed in any cortical or subcortical regions at this threshold.

**Results 2. Stop-signal task performance and behavioral moderation checks**

We examined whether stop-signal task performance, indexed by integrated stop-signal reaction time (iSSRT), could account for moderated ELA-related vulnerability. Poverty-exposed youth showed modestly slower stopping than higher-income youth (mean (SE) iSSRT, 295 (6.26) vs 284 (4.96) ms; Δ = 10.5 ms; *P* = .018). In the comparisons between three groups, iSSRT was similar between the higher-income group and poverty subtype-1, but slightly slower in poverty subtype-2 (mean (SE), 284 (4.97), 290 (6.75), and 300 (6.83) ms for higher-income, poverty subtype-1, and poverty subtype-2, respectively; higher-income vs poverty subtype-2: Δ = 15.6 ms, pFDR = .008; higher-income vs poverty subtype-1: Δ = 5.8 ms, pFDR = .258; poverty subtype-1 vs poverty subtype-2: Δ = 9.8 ms, pFDR = .093; Supplementary Table 5).

In baseline mixed-effects models predicting CBCL total problems, slower iSSRT was associated with higher overall symptom levels (β = 0.048, SE = 0.018; *P* = .0069), but iSSRT did not significantly moderate cumulative-ELA–CBCL coupling (cumulative ELA × iSSRT: β = 0.016, SE = 0.017; *P* = .35), nor did this interaction differ by poverty status (cumulative ELA × iSSRT × poverty: β = −0.062, SE = 0.040; *P* = .12). Thus, in contrast to multivariate neurofunctional patterns, behavioral task performance did not explain variability in vulnerability to cumulative ELA.

**Results 3. Sensitivity analyses**

We conducted three sensitivity analyses to assess robustness of (i) poverty amplification of cumulative-ELA-related vulnerability, (ii) subtype stratification of vulnerability within poverty exposure, and (iii) the synergistic moderation by subtype and inhibitory-control temperament.

**Results 3.1 Sensitivity analysis 1: cumulative ELA defined using all 14 ELAs**

In the primary analyses, cumulative ELA was defined using the eight ELA domains that showed significant poverty × ELA interactions on CBCL total problems in cross-sectional models. To test whether results depended on this selection, we repeated key analyses using cumulative ELA computed from all 14 ELA indicators.

**Poverty × cumulative ELA (14 ELAs) → CBCL total problems.** When cumulative ELA was defined using all 14 indicators, poverty status continued to moderate ELA-related vulnerability. At baseline, the slope linking cumulative ELA (14 ELAs) to CBCL total problems was steeper among children in the poverty-exposed group than among the higher-income group (β_poverty_ (SE) = 0.381 (0.014), β_higher-income_ (SE) = 0.312 (0.004); Δβ (SE) = 0.069 (0.020), *P* < .0001). This group difference remained significant three years later (wave-4: β_poverty_ (SE) = 0.293 (0.015), β_higher-income_ (SE) = 0.247 (0.008); Δβ (SE) = 0.046 (0.017), *P* = .0073). Thus, poverty-related amplification of cumulative-ELA-related vulnerability persisted even when all 14 ELAs were included.

**Subtype × cumulative ELA (14 ELAs) → CBCL total problems.** We next tested whether the two neurofunctional subtypes continued to differ in cumulative-ELA-related vulnerability when using all 14 ELAs. The poverty subtype-1 showed a steeper baseline slope of CBCL total problems on cumulative ELA (14 ELAs) than the higher-income comparison group (β_poverty-subtype-1_ (SE) = 0.413 (0.025), β_higher-income_ (SE) = 0.299 (0.009); Δβ (SE) = 0.113 (0.026), *P* < .0001). For the poverty subtype-2, this difference was not significant (β_poverty-subtype-2_ (SE) = 0.333 (0.025); Δβ (SE) = 0.034 (0.026), *P* = .201). Two years later (wave-3), the slope remained significantly higher for poverty subtype-1 (β_poverty-subtype-1_ (SE) = 0.295 (0.026), β_higher-income_ (SE) = 0.240 (0.009); Δβ (SE) = 0.055 (0.027), *P* = .0413), but not for poverty subtype-2 (β_poverty-subtype-2_ (SE) = 0.283 (0.026); Δβ (SE) = 0.043 (0.028), *P* = .1187). These results indicate that the buffered pattern of cumulative-ELA-related vulnerability in poverty subtype-2 (relative to poverty subtype-1) is robust to an alternative definition of cumulative ELA.

**Subtype × inhibitory-control temperament × cumulative ELA (14 ELAs) → CBCL total problems.** Finally, we tested whether joint moderation by neurofunctional subtype and inhibitory-control temperament remained evident when cumulative ELA included all 14 indicators. The three-way interaction among cumulative ELA, inhibitory-control temperament, and subtype was significant at both follow-ups (wave-3: F = 4.45, *P* = .017; wave-4: F = 3.02, *P* = .049). Simple-slopes analyses showed that, among the higher-income group, the difference in cumulative-ELA-related vulnerability between high and low inhibitory-control groups was small (wave-3: Δβ (SE) = 0.080 (0.047), *P* = .088; wave-4: Δβ (SE) = 0.066 (0.050), *P* = .184). Among the poverty subtype-1, differences were modest and non-significant (wave-3: Δβ (SE) = 0.201 (0.147), *P* = .172; wave-4: Δβ (SE) = 0.251 (0.159), *P* = .114). By contrast, the poverty subtype-2 showed substantially larger differences (wave-3: Δβ (SE) = 0.551 (0.152), *P* < .001; wave-4: Δβ (SE) = 0.466 (0.166), *P* = .005). Notably, among the poverty subtype-2 who also had high inhibitory-control temperament, baseline cumulative ELA (14 ELAs) no longer showed a significant association with CBCL total problems at either follow-up (wave-3: β (SE) = -0.372 (0.209), *P* = .075; wave-4: β (SE) = -0.244 (0.228), *P* = .283). Thus, the synergistic buffering effect of poverty subtype-2 and high inhibitory-control temperament persists when cumulative ELA is defined broadly.

**Results 3.2 Sensitivity analysis 2: cumulative ELA without imputed missing values (complete-case)**

In the primary analyses, missing values for individual ELAs were imputed using mean imputation before computing cumulative ELA. To test whether imputation influenced results, we recomputed cumulative ELA from the eight key ELA indicators using only participants with complete data on all eight indicators, without imputation. Baseline missingness (main sample, N = 10,112) was: parental substance-use problems, 490 (4.8%); experience of trauma, 266 (2.6%); experience of bullying, 2 (0.02%); experience of surgery, 1 (0.01%); parental psychopathology, 54 (0.53%); parental coldness, 0; sleep problems, 202 (2.0%); and prenatal exposure, 1,432 (14.2%). After excluding participants with missing data on any of these eight indicators, 8,094 children remained for complete-case analyses.

**Poverty × cumulative ELA (8 ELAs, complete cases) → CBCL total problems.** At baseline, the poverty-exposed group again showed a steeper slope of CBCL total problems on cumulative ELA (8 ELAs, complete cases) than the higher-income group (β_poverty_ (SE) = 0.481 (0.019), β_higher-income_ (SE) = 0.387 (0.010); Δβ (SE) = 0.094 (0.022), *P* < .0001). This difference remained significant at wave-4 (β_poverty_ (SE) = 0.376 (0.021), β_higher-income_ (SE) = 0.302 (0.011); Δβ (SE) = 0.074 (0.024), *P* = .0019). Thus, the amplification of cumulative-ELA-related vulnerability by poverty status does not depend on imputation.

**Subtype × cumulative ELA (8 ELAs, complete cases) → CBCL total problems.** The poverty subtype-1 again showed heightened vulnerability relative to the higher-income comparison group at baseline (β_poverty-subtype-1_ (SE) = 0.512 (0.034), β_higher-income_ (SE) = 0.377 (0.011); Δβ (SE) = 0.135 (0.036), *P* = .0002). For the poverty subtype-2, the group difference was smaller and not statistically significant (β_poverty-subtype-2_ (SE) = 0.428 (0.035); Δβ (SE) = 0.083 (0.048), *P* = .081). Two years later, the slope remained significantly higher for poverty subtype-1 (wave-3: β_poverty-subtype-1_ (SE) = 0.409 (0.037), β_higher-income_ (SE) = 0.294 (0.012); Δβ (SE) = 0.115 (0.038), *P* = .0028), but not for poverty subtype-2 (β_poverty-subtype-2_ (SE) = 0.363 (0.037); Δβ (SE) = 0.069 (0.039), *P* = .0753). This pattern confirms that the relatively buffered vulnerability in poverty subtype-2 is not an artifact of mean imputation.

**Subtype × inhibitory-control temperament × cumulative ELA (8 ELAs, complete cases) → CBCL total problems.** We repeated the three-way moderation analyses using complete-case cumulative ELA. The interaction among cumulative ELA (8 ELAs), inhibitory-control temperament, and subtype was significant or marginally significant (wave-3: F = 3.97, *P* = .019; wave-4: F = 2.75, *P* = .064). Among the higher-income group, differences in vulnerability between high and low inhibitory-control groups were small (wave-3: Δβ (SE) = 0.050 (0.051), *P* = .328; wave-4: Δβ (SE) = 0.061 (0.054), *P* = .260). Among the poverty subtype-1, differences were modest and non-significant (wave-3: Δβ (SE) = 0.229 (0.164), *P* = .162; wave-4: Δβ (SE) = 0.100 (0.176), *P* = .572). By contrast, the poverty subtype-2 again showed larger differences (wave-3: Δβ (SE) = 0.501 (0.158), *P* = .002; wave-4: Δβ (SE) = 0.485 (0.174), *P* = .005). Critically, among the poverty subtype-2 who also had high inhibitory-control temperament, baseline cumulative ELA (8 ELAs, complete cases) did not significantly predict CBCL total problems at either wave (wave-3: β (SE) = -0.301 (0.213), *P* = .157; wave-4: β (SE) = -0.276 (0.233), *P* = .236). These results indicate that the buffered vulnerability associated with the combination of poverty subtype-2 and high inhibitory-control temperament is robust to the handling of missing ELA data.

**Results 3.3 Sensitivity analysis 3: cumulative ELA after random sibling exclusion**

The ABCD Study oversampled twins and siblings.^9^ In our main sample (N = 10,112), 1,700 children (16.8%) had at least one sibling also enrolled. Therefore, we conducted an additional sensitivity analysis that removed sibling clusters by randomly retaining only one child per family. After randomly selecting one sibling per family for all families with more than one participating child, 8,414 children remained at baseline.

**Poverty × cumulative ELA → CBCL total problems (reduced dataset).** In this reduced sample, poverty status continued to moderate cumulative-ELA-related vulnerability. At baseline, the slope of CBCL total problems on cumulative ELA (8 ELAs used in the main cumulative index) was steeper in the poverty-exposed group (β_poverty_ (SE) = 0.497 (0.019), β_higher-income_ (SE) = 0.410 (0.010); Δβ (SE) = 0.087 (0.021), *P* < .0001).This difference remained significant at wave-4 (β_poverty_ (SE) = 0.404 (0.021), β_higher-income_ (SE) = 0.335 (0.010); Δβ (SE) = 0.068 (0.023), *P* = .0034). Thus, the amplification of cumulative-ELA-related vulnerability by poverty status is not driven by sibling clustering.

**Subtype × cumulative ELA → CBCL total problems (reduced dataset).** In the reduced dataset, the poverty subtype-1 continued to show heightened vulnerability relative to the higher-income comparison group at baseline (β_poverty-subtype-1_ (SE) = 0.548 (0.034), β_higher-income_ (SE) = 0.391 (0.011); Δβ (SE) = 0.157 (0.036), *P* < .0001). The poverty subtype-2 again showed a smaller and non-significant difference (β_poverty-subtype-2_ (SE) = 0.452 (0.036); Δβ (SE) = 0.061 (0.038), *P* = .108). At wave-3, the slope remained significantly higher for poverty subtype-1 (β_poverty-subtype-1_ (SE) = 0.422 (0.036), β_higher-income_ (SE) = 0.316 (0.012); Δβ (SE) = 0.106 (0.038), *P* = .0055), but not for poverty subtype-2 (β_poverty-subtype-2_ (SE) = 0.393 (0.039); Δβ (SE) = 0.077 (0.041), *P* = .0578). This supports the conclusion that the buffered vulnerability in poverty subtype-2 is not explained by sibling structure.

**Subtype × inhibitory-control temperament × cumulative ELA → CBCL total problems (reduced dataset).** We repeated the three-way moderation analysis in the reduced sample. The interaction among cumulative ELA, inhibitory-control temperament, and subtype was significant at wave-3 and non-significant at wave-4 (wave-3: F = 4.14, *P* = .016; wave-4: F = 1.33, *P* = .266). Among the higher-income group, differences in vulnerability between high and low inhibitory-control groups remained modest (wave-3: Δβ (SE) = 0.098 (0.051), *P* = .053; wave-4: Δβ (SE) = 0.098 (0.054), *P* = .072). Among the poverty subtype-1, differences were modest and non-significant (wave-3: Δβ (SE) = 0.184 (0.153), *P* = .229; wave-4: Δβ (SE) = 0.115 (0.164), *P* = .482). The poverty subtype-2 again showed substantially larger differences (wave-3: Δβ (SE) = 0.615 (0.173), *P* < .001; wave-4: Δβ (SE) = 0.417 (0.189), *P* = .027). In addition, among the poverty subtype-2 who also had high inhibitory-control temperament, baseline cumulative ELA did not significantly predict CBCL total problems at either follow-up (wave-3: β (SE) = -0.462 (0.239), *P* = .053; wave-4: β (SE) = -0.158 (0.260), *P* = .545). These findings indicate that the buffered cumulative-ELA-related vulnerability associated with the combined neurofunctional and temperamental profile in poverty subtype-2 is not attributable to sibling-related family clustering.

**Results 4. Multiscale molecular correspondence**

**Neurotransmitter receptor/transporter correspondence.** We first tested whether subtype-specific subtype–higher-income β difference maps (standardized coefficients) aligned with PET-derived neurotransmitter receptor/transporter distributions using spin-based inference (10,000 rotations). The high-activation poverty subtype-1 showed significant positive correspondence with monoaminergic receptor topographies, including serotonergic 5-HT1A (r = .202; P_spin_ = .048) and dopaminergic D2 distributions (D2_1_: r = .177, P_spin_ = .050; D2_3_: r = .209, P_spin_ = .042). These correspondences are summarized in Figure 4F and illustrated in Figure 4G; full results are reported in Supplementary Table 9. In contrast, the low-activation poverty subtype-2 showed no significant receptor/transporter correspondence after accounting for spatial autocorrelation (Figure 4F; Supplementary Table 9).

**Transcriptomic correspondence.** We next evaluated transcriptional correspondence using AHBA cortical expression profiles (15,632 genes) and spin-based inference. PLS1 showed significant covariance with the subtype-2–higher-income β difference map (standardized coefficients; variance explained = 38.6%; P_spin_ = .024) but not with the subtype-1–higher-income β difference map (30.7%; P_spin_ = .233), indicating a subtype-specific transcriptomic signature (Figure 4B). For poverty subtype-2, genes with strong negative PLS1 loadings (e.g., NAT8L, near the bottom of the PLS1 weight distribution) were more highly expressed in regions showing larger negative subtype-2–higher-income β differences (examples in Figure 4C; spatial correspondence shown in Figure 4D). Cell-type enrichment analysis (Seidlitz et al., 2020) revealed that genes associated with the poverty subtype-2 β difference pattern were significantly enriched in astrocytes (median rank = 3745 / 15632, *P* < .0001), oligodendrocytes (median rank = 4014 / 15632, *P* < .0001), microglia (median rank = 5565 / 15632, *P* < .0001), and oligodendroglial precursor cells (OPCs; median rank = 6480 / 15632, *P* < .0001), and underrepresented in excitatory neurons (median rank = 10312 / 15632, *P* < .0001), inhibitory neurons (median rank = 9195.5 / 15632, *P* < .0001), and endothelial cells (median rank = 8359.5 / 15632, P = .0189; Figure 4E). Together, these results form a coherent divergence pattern: poverty subtype-1 aligns with serotonergic/dopaminergic receptor distributions, whereas poverty subtype-2 is characterized primarily by a transcriptomic signature enriched in glial and precursor cell classes.

**Supplementary Tables**

**Table 1. Associations between each early-life adversity (ELA) indicator and CBCL total problems (ELA–CBCL coupling) by poverty and interaction tests**

| ELA indicator | | Poverty × ELA interaction, β (95% CI) | Poverty × ELA interaction, *P* value | Poverty × ELA interaction, *P*_FDR_ value | Poverty-exposed, β (95% CI) | Higher-income, β (95% CI) |
| --- | --- | --- | --- | --- | --- | --- |
| Experience of surgery | | 0.208 (0.071 to 0.344) | .003 | .008 | 0.343 (0.214 to 0.471) | 0.135 (0.088 to 0.182) |
| Prenatal exposure | | 0.176 (0.123 to 0.229) | <.0001 | <.0001 | 0.348 (0.300 to 0.396) | 0.172 (0.149 to 0.195) |
| Experience of trauma | | 0.278 (0.169 to 0.386) | <.0001 | <.0001 | 0.605 (0.506 to 0.704) | 0.327 (0.283 to 0.371) |
| Parent has substance abuse problem | | 0.204 (0.075 to 0.332) | .002 | .007 | 0.507 (0.393 to 0.622) | 0.304 (0.245 to 0.362) |
| Parental separation | | 0.086 (-0.036 to 0.209) | .165 | .231 | 0.230 (0.121 to 0.340) | 0.144 (0.085 to 0.203) |
| Area deprivation index | | 0.031 (-0.027 to 0.088) | .300 | .35 | 0.094 (0.038 to 0.149) | 0.063 (0.031 to 0.096) |
| Neighborhood unsafety | | 0.046 (-0.004 to 0.096) | .071 | .111 | 0.163 (0.119 to 0.207) | 0.117 (0.092 to 0.142) |
| Sleep problem | | 0.085 (0.034 to 0.135) | .001 | .004 | 0.384 (0.338 to 0.431) | 0.300 (0.281 to 0.319) |
| Experience of being bullied | | 0.167 (0.042 to 0.291) | .009 | .02 | 0.876 (0.764 to 0.987) | 0.709 (0.653 to 0.765) |
| Adverse school environment | | 0.024 (-0.021 to 0.070) | .294 | .35 | 0.119 (0.079 to 0.160) | 0.095 (0.075 to 0.116) |
| Family conflict | | 0.052 (0.002 to 0.103) | .041 | .07 | 0.339 (0.294 to 0.385) | 0.287 (0.266 to 0.308) |
| Parent psychopathology | | 0.051 (0.011 to 0.091) | .012 | .023 | 0.617 (0.582 to 0.653) | 0.566 (0.548 to 0.585) |
| Parent neglect | | 0.000 (-0.044 to 0.045) | .992 | .999 | 0.100 (0.061 to 0.140) | 0.100 (0.079 to 0.121) |
| Parent coldness | | -0.000 (-0.048 to 0.048) | .999 | .999 | 0.073 (0.029 to 0.117) | 0.073 (0.052 to 0.093) |
|  | Abbreviations: CBCL, Child Behavior Checklist; CI, confidence interval; ELA, early-life adversity. β values are standardized regression coefficients. The interaction term corresponds to poverty status × ELA (poverty moderation of each ELA–CBCL association), estimated from the models described in Methods.  P values are from poverty × ELA interaction terms. *P*_FDR_ values are Benjamini-Hochberg adjusted across the 14 ELA indicators.  The primary 8-ELA index was defined as the subset of ELAs with **nominal** poverty×ELA interaction P < .05 in ELA-specific models; 7 of these 8 interactions survived FDR correction (*P*_FDR_ < .05) | | | | | |

**Table 2. Standardized associations between cumulative ELA and CBCL total problems (cumulative ELA–CBCL coupling) across waves, stratified by poverty status (before fMRI QC) and neurofunctional subtype (after fMRI QC)**

| CBCL total problems ~ cumulative ELA | β (95% CI) | | | | |
| --- | --- | --- | --- | --- | --- |
|  | Before fMRI QC | | After fMRI QC | | |
|  | Higher-income group | Poverty-exposed group | Higher-income group | Poverty subtype-1 | Poverty subtype-2 |
| CBCL total problems at wave-1 (baseline) | 0.388 (0.369, 0.406) | 0.468 (0.432, 0.504) | 0.376 (0.355, 0.397) | 0.516 (0.452, 0.580) | 0.409 (0.346, 0.473) |
| CBCL total problems at wave-2 | 0.334 (0.315, 0.352) | 0.413 (0.376, 0.449) | 0.325 (0.304, 0.347) | 0.436 (0.370, 0.501) | 0.401 (0.336, 0.466) |
| CBCL total problems at wave-3 | 0.320 (0.301, 0.338) | 0.396 (0.359, 0.434) | 0.306 (0.284, 0.327) | 0.417 (0.350, 0.483) | 0.375 (0.309, 0.440) |
| CBCL total problems at wave-4 | 0.319 (0.300, 0.338) | 0.380 (0.341, 0.418) | 0.318 (0.296, 0.340) | 0.386 (0.318, 0.455) | 0.380 (0.312, 0.449) |
| Abbreviations: CBCL, Child Behavior Checklist; CI, confidence interval; ELA, early-life adversity; QC, quality control. Entries are standardized regression coefficients (β) and 95% CIs for the association between cumulative ELA burden and CBCL total problems at each wave, based on the models described in Methods. “Before task-fMRI QC” refers to the behavioral analytic cohort; “After task-fMRI QC” refers to the subset with usable stop-signal task fMRI data after QC (see Supplementary Methods 1). | | | | | |

**Table 3. Differences in cumulative ELA–CBCL coupling between the poverty-exposed and higher-income groups across waves**

| Δβ (95% CI) | | | |
| --- | --- | --- | --- |
| Wave-1 (baseline) | Wave-2 | Wave-3 | Wave-4 |
| 0.080 (0.040 to 0.120) *** | 0.079 (0.038 to 0.12) *** | 0.077 (0.035 to 0.118) *** | 0.061 (0.018 to 0.104) ** |
| Entries show the difference in standardized cumulative ELA–CBCL slopes (Δβ = β_poverty-exposed_ – β_higher-income_) at each wave, based on the models described in Methods.  ***, **: *P* < .001, .01 after false discovery rate (FDR) correction across time-point comparisons. | | | |

**Table 4. Regional differences in stop-signal task activation between poverty subtypes and the higher-income group**

| Brain Region | Poverty subtype-1 vs higher-income group, β (95% CI) | Poverty subtype-2 vs higher-income group, β (95% CI) |
| --- | --- | --- |
| 3rd-ventricle | 0.09 (-0.03 to 0.21) | -0.22 (-0.34 to -0.09) * |
| 4th-ventricle | 0.09 (-0.03 to 0.22) | -0.11 (-0.24 to 0.01) |
| L-Accumbens-area | 0.25 (0.13 to 0.38) * | -0.34 (-0.47 to -0.22) * |
| R-Accumbens-area | 0.18 (0.06 to 0.31) * | -0.40 (-0.52 to -0.27) * |
| L-Amygdala | 0.28 (0.16 to 0.41) * | -0.31 (-0.44 to -0.19) * |
| R-Amygdala | 0.18 (0.05 to 0.30) * | -0.38 (-0.50 to -0.25) * |
| Brain-Stem | 0.30 (0.18 to 0.42) * | -0.32 (-0.45 to -0.20) * |
| L-Cerebellum-White-Matter | 0.42 (0.29 to 0.54) * | -0.54 (-0.67 to -0.42) * |
| R-Cerebellum-White-Matter | 0.47 (0.35 to 0.59) * | -0.56 (-0.68 to -0.43) * |
| L-Caudate | 0.40 (0.28 to 0.52) * | -0.58 (-0.70 to -0.45) * |
| R-Caudate | 0.42 (0.30 to 0.54) * | -0.54 (-0.66 to -0.41) * |
| L-Cerebellum-Cortex | 0.31 (0.18 to 0.43) * | -0.34 (-0.47 to -0.22) * |
| R-Cerebellum-Cortex | 0.41 (0.28 to 0.53) * | -0.36 (-0.48 to -0.23) * |
| L-Cerebellum-White-Matter | 0.22 (0.09 to 0.34) * | -0.33 (-0.45 to -0.20) * |
| R-Cerebellum-White-Matter | 0.33 (0.21 to 0.45) * | -0.32 (-0.44 to -0.19) * |
| csf | 0.37 (0.25 to 0.50) * | -0.39 (-0.52 to -0.27) * |
| L-Hippocampus | 0.29 (0.17 to 0.42) * | -0.43 (-0.55 to -0.30) * |
| R-Hippocampus | 0.27 (0.15 to 0.39) * | -0.49 (-0.62 to -0.37) * |
| L-Inf-Lat-Vent | 0.02 (-0.11 to 0.14) | -0.06 (-0.19 to 0.06) |
| R-Inf-Lat-Vent | 0.01 (-0.11 to 0.14) | -0.07 (-0.20 to 0.05) |
| L-Lateral-Ventricle | 0.12 (-0.01 to 0.24) | -0.11 (-0.24 to 0.02) |
| R-Lateral-Ventricle | 0.13 (0.01 to 0.26) * | -0.15 (-0.27 to -0.02) * |
| L-Pallidum | 0.02 (-0.10 to 0.15) | -0.36 (-0.49 to -0.23) * |
| R-Pallidum | 0.20 (0.07 to 0.32) * | -0.26 (-0.39 to -0.14) * |
| L-Putamen | 0.29 (0.17 to 0.42) * | -0.56 (-0.68 to -0.43) * |
| R-Putamen | 0.35 (0.22 to 0.47) * | -0.50 (-0.63 to -0.38) * |
| L-Thalamus-Proper | 0.40 (0.28 to 0.52) * | -0.55 (-0.67 to -0.42) * |
| R-Thalamus-Proper | 0.39 (0.27 to 0.51) * | -0.54 (-0.66 to -0.41) * |
| L-VentralDC | 0.34 (0.22 to 0.46) * | -0.42 (-0.55 to -0.30) * |
| R-VentralDC | 0.29 (0.17 to 0.41) * | -0.44 (-0.56 to -0.31) * |
| L-G and S frontomargin | 0.32 (0.20 to 0.45) * | -0.34 (-0.47 to -0.22) * |
| L-G and S occipital inf | 0.37 (0.24 to 0.49) * | -0.49 (-0.61 to -0.36) * |
| L-G and S paracentral | 0.32 (0.20 to 0.44) * | -0.56 (-0.68 to -0.43) * |
| L-G and S subcentral | 0.22 (0.09 to 0.34) * | -0.45 (-0.57 to -0.32) * |
| L-G and S transv frontopol | 0.23 (0.11 to 0.36) * | -0.29 (-0.41 to -0.16) * |
| L-G and S cingul-Ant | 0.30 (0.17 to 0.42) * | -0.43 (-0.56 to -0.31) * |
| L-G and S cingul-Mid-Ant | 0.43 (0.31 to 0.55) * | -0.57 (-0.70 to -0.45) * |
| L-G and S cingul-Mid-Post | 0.38 (0.26 to 0.50) * | -0.56 (-0.68 to -0.43) * |
| L-G cingul-Post-dorsal | 0.31 (0.19 to 0.43) * | -0.51 (-0.63 to -0.38) * |
| L-G cingul-Post-ventral | 0.34 (0.22 to 0.46) * | -0.50 (-0.63 to -0.38) * |
| L-G cuneus | 0.40 (0.27 to 0.52) * | -0.50 (-0.62 to -0.37) * |
| L-G front inf-Opercular | 0.36 (0.24 to 0.48) * | -0.53 (-0.66 to -0.41) * |
| L-G front inf-Orbital | 0.18 (0.06 to 0.30) * | -0.30 (-0.43 to -0.18) * |
| L-G front inf-Triangul | 0.20 (0.08 to 0.33) * | -0.34 (-0.46 to -0.21) * |
| L-G front middle | 0.46 (0.33 to 0.58) * | -0.56 (-0.68 to -0.44) * |
| L-G front sup | 0.37 (0.25 to 0.49) * | -0.52 (-0.64 to -0.39) * |
| L-G Ins lg and S cent ins | 0.33 (0.21 to 0.46) * | -0.42 (-0.55 to -0.30) * |
| L-G insular short | 0.39 (0.27 to 0.51) * | -0.46 (-0.59 to -0.34) * |
| L-G occipital middle | 0.43 (0.31 to 0.55) * | -0.50 (-0.62 to -0.37) * |
| L-G occipital sup | 0.45 (0.32 to 0.57) * | -0.44 (-0.56 to -0.31) * |
| L-G oc-temp lat-fusifor | 0.41 (0.29 to 0.53) * | -0.57 (-0.69 to -0.44) * |
| L-G oc-temp med-Lingual | 0.45 (0.33 to 0.57) * | -0.50 (-0.63 to -0.38) * |
| L-G oc-temp med-Parahip | 0.23 (0.11 to 0.35) * | -0.51 (-0.64 to -0.39) * |
| L-G orbital | 0.27 (0.14 to 0.39) * | -0.35 (-0.48 to -0.23) * |
| L-G pariet inf-Angular | 0.36 (0.24 to 0.48) * | -0.56 (-0.69 to -0.44) * |
| L-G pariet inf-Supramar | 0.40 (0.28 to 0.52) * | -0.54 (-0.67 to -0.42) * |
| L-G parietal sup | 0.49 (0.37 to 0.61) * | -0.53 (-0.65 to -0.40) * |
| L-G postcentral | 0.38 (0.26 to 0.51) * | -0.48 (-0.60 to -0.35) * |
| L-G precentral | 0.39 (0.27 to 0.52) * | -0.61 (-0.74 to -0.49) * |
| L-G precuneus | 0.40 (0.28 to 0.53) * | -0.47 (-0.59 to -0.34) * |
| L-G rectus | 0.15 (0.03 to 0.28) * | -0.24 (-0.36 to -0.11) * |
| L-G subcallosal | 0.18 (0.05 to 0.30) * | -0.30 (-0.43 to -0.17) * |
| L-G temp sup-G T transv | 0.20 (0.08 to 0.32) * | -0.42 (-0.55 to -0.30) * |
| L-G temp sup-Lateral | 0.22 (0.10 to 0.35) * | -0.49 (-0.62 to -0.37) * |
| L-G temp sup-Plan polar | 0.25 (0.13 to 0.38) * | -0.27 (-0.39 to -0.14) * |
| L-G temp sup-Plan tempo | 0.33 (0.21 to 0.46) * | -0.57 (-0.70 to -0.45) * |
| L-G temporal inf | 0.19 (0.06 to 0.31) * | -0.34 (-0.47 to -0.21) * |
| L-G temporal middle | 0.18 (0.06 to 0.31) * | -0.48 (-0.61 to -0.35) * |
| L-Lat Fis-ant-Horizont | 0.19 (0.07 to 0.31) * | -0.30 (-0.43 to -0.18) * |
| L-Lat Fis-ant-Vertical | 0.18 (0.05 to 0.30) * | -0.36 (-0.49 to -0.23) * |
| L-Lat Fis-post | 0.31 (0.19 to 0.44) * | -0.52 (-0.65 to -0.40) * |
| L-Pole occipital | 0.27 (0.14 to 0.39) * | -0.18 (-0.31 to -0.06) * |
| L-Pole temporal | 0.11 (-0.01 to 0.24) | -0.28 (-0.41 to -0.15) * |
| L-S calcarine | 0.42 (0.30 to 0.55) * | -0.55 (-0.67 to -0.42) * |
| L-S central | 0.37 (0.25 to 0.49) * | -0.52 (-0.65 to -0.40) * |
| L-S cingul-Marginalis | 0.45 (0.33 to 0.57) * | -0.56 (-0.68 to -0.43) * |
| L-S circular insula ant | 0.33 (0.20 to 0.45) * | -0.46 (-0.59 to -0.34) * |
| L-S circular insula inf | 0.20 (0.08 to 0.33) * | -0.49 (-0.62 to -0.37) * |
| L-S circular insula sup | 0.37 (0.24 to 0.49) * | -0.47 (-0.60 to -0.35) * |
| L-S collat transv ant | 0.29 (0.17 to 0.41) * | -0.31 (-0.44 to -0.19) * |
| L-S collat transv post | 0.42 (0.30 to 0.54) * | -0.32 (-0.44 to -0.19) * |
| L-S front inf | 0.45 (0.33 to 0.58) * | -0.53 (-0.65 to -0.40) * |
| L-S front middle | 0.41 (0.29 to 0.53) * | -0.41 (-0.53 to -0.28) * |
| L-S front sup | 0.40 (0.28 to 0.53) * | -0.49 (-0.62 to -0.37) * |
| L-S interm prim-Jensen | 0.31 (0.18 to 0.43) * | -0.47 (-0.59 to -0.34) * |
| L-S intrapariet and P trans | 0.49 (0.37 to 0.61) * | -0.57 (-0.69 to -0.44) * |
| L-S oc middle and Lunatus | 0.38 (0.25 to 0.50) * | -0.40 (-0.53 to -0.28) * |
| L-S oc sup and transversal | 0.36 (0.24 to 0.49) * | -0.47 (-0.59 to -0.34) * |
| L-S occipital ant | 0.36 (0.23 to 0.48) * | -0.50 (-0.62 to -0.37) * |
| L-S oc-temp lat | 0.31 (0.19 to 0.44) * | -0.57 (-0.70 to -0.45) * |
| L-S oc-temp med and Lingual | 0.35 (0.23 to 0.48) * | -0.42 (-0.55 to -0.30) * |
| L-S orbital lateral | 0.19 (0.07 to 0.32) * | -0.25 (-0.38 to -0.12) * |
| L-S orbital med-olfact | 0.20 (0.08 to 0.33) * | -0.18 (-0.31 to -0.06) * |
| L-S orbital-H Shaped | 0.31 (0.19 to 0.44) * | -0.50 (-0.63 to -0.38) * |
| L-S parieto occipital | 0.35 (0.23 to 0.48) * | -0.52 (-0.64 to -0.39) * |
| L-S pericallosal | 0.44 (0.32 to 0.56) * | -0.56 (-0.68 to -0.44) * |
| L-S postcentral | 0.36 (0.24 to 0.49) * | -0.52 (-0.64 to -0.39) * |
| L-S precentral-inf-part | 0.37 (0.25 to 0.50) * | -0.58 (-0.70 to -0.45) * |
| L-S precentral-sup-part | 0.39 (0.27 to 0.51) * | -0.54 (-0.67 to -0.42) * |
| L-S suborbital | 0.26 (0.13 to 0.38) * | -0.39 (-0.52 to -0.27) * |
| L-S subparietal | 0.18 (0.06 to 0.31) * | -0.42 (-0.55 to -0.30) * |
| L-S temporal inf | 0.24 (0.11 to 0.36) * | -0.40 (-0.53 to -0.28) * |
| L-S temporal sup | 0.24 (0.11 to 0.36) * | -0.54 (-0.67 to -0.42) * |
| L-S temporal transverse | 0.25 (0.13 to 0.38) * | -0.39 (-0.51 to -0.26) * |
| R-G and S frontomargin | 0.25 (0.13 to 0.37) * | -0.48 (-0.61 to -0.35) * |
| R-G and S occipital inf | 0.46 (0.34 to 0.59) * | -0.52 (-0.64 to -0.39) * |
| R-G and S paracentral | 0.33 (0.21 to 0.45) * | -0.50 (-0.63 to -0.37) * |
| R-G and S subcentral | 0.30 (0.18 to 0.42) * | -0.47 (-0.59 to -0.34) * |
| R-G and S transv frontopol | 0.25 (0.13 to 0.38) * | -0.31 (-0.43 to -0.18) * |
| R-G and S cingul-Ant | 0.31 (0.19 to 0.43) * | -0.48 (-0.61 to -0.36) * |
| R-G and S cingul-Mid-Ant | 0.41 (0.29 to 0.53) * | -0.50 (-0.63 to -0.38) * |
| R-G and S cingul-Mid-Post | 0.40 (0.28 to 0.52) * | -0.57 (-0.70 to -0.45) * |
| R-G cingul-Post-dorsal | 0.36 (0.24 to 0.48) * | -0.50 (-0.63 to -0.38) * |
| R-G cingul-Post-ventral | 0.40 (0.28 to 0.53) * | -0.49 (-0.62 to -0.37) * |
| R-G cuneus | 0.44 (0.32 to 0.56) * | -0.48 (-0.60 to -0.35) * |
| R-G front inf-Opercular | 0.41 (0.29 to 0.53) * | -0.55 (-0.67 to -0.42) * |
| R-G front inf-Orbital | 0.36 (0.23 to 0.48) * | -0.40 (-0.53 to -0.28) * |
| R-G front inf-Triangul | 0.37 (0.25 to 0.50) * | -0.51 (-0.64 to -0.39) * |
| R-G front middle | 0.53 (0.41 to 0.65) * | -0.58 (-0.70 to -0.46) * |
| R-G front sup | 0.43 (0.31 to 0.55) * | -0.53 (-0.65 to -0.40) * |
| R-G Ins lg and S cent ins | 0.40 (0.27 to 0.52) * | -0.39 (-0.51 to -0.26) * |
| R-G insular short | 0.31 (0.18 to 0.43) * | -0.46 (-0.59 to -0.34) * |
| R-G occipital middle | 0.44 (0.31 to 0.56) * | -0.55 (-0.68 to -0.43) * |
| R-G occipital sup | 0.38 (0.26 to 0.51) * | -0.44 (-0.57 to -0.32) * |
| R-G oc-temp lat-fusifor | 0.45 (0.32 to 0.57) * | -0.55 (-0.68 to -0.43) * |
| R-G oc-temp med-Lingual | 0.50 (0.38 to 0.62) * | -0.50 (-0.62 to -0.37) * |
| R-G oc-temp med-Parahip | 0.14 (0.01 to 0.26) * | -0.42 (-0.55 to -0.30) * |
| R-G orbital | 0.30 (0.18 to 0.42) * | -0.35 (-0.48 to -0.22) * |
| R-G pariet inf-Angular | 0.36 (0.24 to 0.48) * | -0.56 (-0.69 to -0.44) * |
| R-G pariet inf-Supramar | 0.47 (0.35 to 0.59) * | -0.54 (-0.66 to -0.41) * |
| R-G parietal sup | 0.54 (0.41 to 0.66) * | -0.53 (-0.65 to -0.40) * |
| R-G postcentral | 0.38 (0.26 to 0.50) * | -0.51 (-0.64 to -0.39) * |
| R-G precentral | 0.46 (0.34 to 0.59) * | -0.60 (-0.73 to -0.48) * |
| R-G precuneus | 0.39 (0.27 to 0.52) * | -0.51 (-0.64 to -0.39) * |
| R-G rectus | 0.12 (-0.00 to 0.24) | -0.21 (-0.34 to -0.09) * |
| R-G subcallosal | 0.23 (0.11 to 0.35) * | -0.31 (-0.43 to -0.18) * |
| R-G temp sup-G T transv | 0.28 (0.16 to 0.40) * | -0.45 (-0.58 to -0.33) * |
| R-G temp sup-Lateral | 0.39 (0.27 to 0.51) * | -0.58 (-0.71 to -0.46) * |
| R-G temp sup-Plan polar | 0.28 (0.16 to 0.40) * | -0.36 (-0.49 to -0.24) * |
| R-G temp sup-Plan tempo | 0.32 (0.20 to 0.45) * | -0.49 (-0.62 to -0.37) * |
| R-G temporal inf | 0.08 (-0.04 to 0.21) | -0.44 (-0.56 to -0.31) * |
| R-G temporal middle | 0.32 (0.20 to 0.45) * | -0.56 (-0.68 to -0.43) * |
| R-Lat Fis-ant-Horizont | 0.23 (0.11 to 0.36) * | -0.43 (-0.55 to -0.30) * |
| R-Lat Fis-ant-Vertical | 0.21 (0.08 to 0.33) * | -0.32 (-0.45 to -0.20) * |
| R-Lat Fis-post | 0.33 (0.21 to 0.46) * | -0.50 (-0.62 to -0.37) * |
| R-Pole occipital | 0.34 (0.22 to 0.46) * | -0.41 (-0.54 to -0.28) * |
| R-Pole temporal | 0.21 (0.09 to 0.34) * | -0.32 (-0.45 to -0.19) * |
| R-S calcarine | 0.39 (0.27 to 0.52) * | -0.52 (-0.64 to -0.39) * |
| R-S central | 0.35 (0.23 to 0.48) * | -0.53 (-0.66 to -0.41) * |
| R-S cingul-Marginalis | 0.44 (0.32 to 0.56) * | -0.58 (-0.71 to -0.46) * |
| R-S circular insula ant | 0.26 (0.14 to 0.38) * | -0.32 (-0.45 to -0.20) * |
| R-S circular insula inf | 0.20 (0.08 to 0.32) * | -0.45 (-0.58 to -0.33) * |
| R-S circular insula sup | 0.37 (0.24 to 0.49) * | -0.49 (-0.61 to -0.36) * |
| R-S collat transv ant | 0.21 (0.08 to 0.33) * | -0.35 (-0.47 to -0.22) * |
| R-S collat transv post | 0.30 (0.17 to 0.42) * | -0.36 (-0.49 to -0.23) * |
| R-S front inf | 0.48 (0.36 to 0.60) * | -0.60 (-0.72 to -0.47) * |
| R-S front middle | 0.45 (0.32 to 0.57) * | -0.42 (-0.54 to -0.29) * |
| R-S front sup | 0.44 (0.32 to 0.56) * | -0.52 (-0.65 to -0.40) * |
| R-S interm prim-Jensen | 0.23 (0.11 to 0.35) * | -0.54 (-0.66 to -0.41) * |
| R-S intrapariet and P trans | 0.47 (0.35 to 0.59) * | -0.58 (-0.71 to -0.46) * |
| R-S oc middle and Lunatus | 0.42 (0.30 to 0.55) * | -0.45 (-0.58 to -0.33) * |
| R-S oc sup and transversal | 0.40 (0.28 to 0.53) * | -0.53 (-0.66 to -0.41) * |
| R-S occipital ant | 0.44 (0.32 to 0.57) * | -0.56 (-0.69 to -0.44) * |
| R-S oc-temp lat | 0.40 (0.28 to 0.52) * | -0.40 (-0.53 to -0.28) * |
| R-S oc-temp med and Lingual | 0.42 (0.30 to 0.54) * | -0.46 (-0.58 to -0.33) * |
| R-S orbital lateral | 0.33 (0.20 to 0.45) * | -0.39 (-0.51 to -0.26) * |
| R-S orbital med-olfact | 0.17 (0.05 to 0.30) * | -0.30 (-0.43 to -0.17) * |
| R-S orbital-H Shaped | 0.34 (0.21 to 0.46) * | -0.52 (-0.65 to -0.39) * |
| R-S parieto occipital | 0.35 (0.23 to 0.48) * | -0.51 (-0.64 to -0.39) * |
| R-S pericallosal | 0.42 (0.30 to 0.54) * | -0.56 (-0.68 to -0.43) * |
| R-S postcentral | 0.42 (0.30 to 0.55) * | -0.58 (-0.71 to -0.46) * |
| R-S precentral-inf-part | 0.44 (0.32 to 0.57) * | -0.56 (-0.68 to -0.43) * |
| R-S precentral-sup-part | 0.40 (0.27 to 0.52) * | -0.56 (-0.69 to -0.44) * |
| R-S suborbital | 0.22 (0.09 to 0.34) * | -0.38 (-0.50 to -0.25) * |
| R-S subparietal | 0.28 (0.16 to 0.40) * | -0.45 (-0.57 to -0.32) * |
| R-S temporal inf | 0.13 (0.00 to 0.25) * | -0.35 (-0.47 to -0.22) * |
| R-S temporal sup | 0.31 (0.18 to 0.43) * | -0.63 (-0.75 to -0.50) * |
| R-S temporal transverse | 0.32 (0.20 to 0.44) * | -0.46 (-0.59 to -0.34) * |

**Table 5. Participant characteristics across the higher-income group and poverty subtypes in the task-fMRI analytic sample**

|  | Mean (SD) or No. (%) | | | P value (subtype-1 vs subtype-2) |
| --- | --- | --- | --- | --- |
|  | Higher-income group | Poverty subtype-1 | Poverty subtype-2 |  |
| Sex, female, No. (%) | 3212 (49.6) | 235 (48.9) | 225 (51.3) | >.05 |
| Race and Ethnicity (%) White/Black/Hispanic/Asian/Other | 64/7.5/16.1/2.2/10.2 | 17.7/35.3/34.7/0.8/11.4* | 21.6/31/36.9/0.7/9.8* | >.05 |
| Age at baseline, mean (SD), y | 9.95 (0.63) | 9.93 (0.61) | 9.89 (0.63) | >.05 |
| Log (Income to needs ratio), M (SD) | 5.96 (0.63) | 3.61 (0.82) * | 3.62 (0.79) * | >.05 |
| CBCL total problems, Mean (SD) | 44.97 (10.60) | 46.90 (13.03) * | 46.93 (12.12) * | >.05 |
| Parent has substance abuse problem, No., (%) | 1047 (16.9) | 132 (29.7) * | 122 (29.6) * | >.05 |
| Experience of trauma, No., (%) | 2136 (33.7) | 203 (44.0) * | 187 (43.8) * | >.05 |
| Parental separation, No., (%) | 1090 (16.8) | 296 (62.6) * | 249 (57.2) * | >.05 |
| Experience of being bullied, No., (%) | 834 (12.9) | 83 (17.3) * | 89 (20.3) * | >.05 |
| Experience of surgery, No., (%) | 1494 (23.0) | 82 (17.0) | 75 (17.1) | >.05 |
| Parent psychopathology, Mean (SD) | 42.72 (9.62) | 44.40 (12.10) * | 44.21 (11.58) * | >.05 |
| Parent coldness, Mean (SD) | 1.21 (0.29) | 1.25 (0.34) * | 1.25 (0.33) * | >.05 |
| Family conflict, Mean (SD) | 2.45 (1.93) | 2.91 (2.11) * | 2.69 (2.06) * | >.05 |
| Parent neglect, Mean (SD) | 1.57 (0.46) | 1.69 (0.60) * | 1.72 (0.61) * | >.05 |
| Sleep problem, Mean (SD) | 0.44 (0.76) | 0.38 (0.71) | 0.38 (0.75) | >.05 |
| Prenatal exposure, Mean (SD) | 1.73 (1.64) | 2.19 (1.79) * | 2.21 (1.89) * | >.05 |
| Area deprivation index, Mean (SD) | 34.36 (23.22) | 59.43 (28.70) * | 58.46 (27.40) * | >.05 |
| Neighborhood unsafety, Mean (SD) | 1.94 (0.86) | 2.70 (1.14) * | 2.66 (1.14) * | >.05 |
| Adverse school environment, Mean (SD) | 10.06 (2.66) | 9.96 (3.16) | 9.91 (3.06) | >.05 |
| Cumulative ELAs-8 sig vars, M (SD) | 1.63 (1.05) | 1.92 (1.22) * | 1.92 (1.29) * | >.05 |
| Cumulative ELAs-14 vars, Mean (SD) | 2.84 (1.36) | 4.05 (1.60) * | 3.99 (1.63) * | >.05 |
| SST-reaction time, Mean (SD), ms | 284 (4.97) | 290 (6.75) | 300 (6.83) * | >.05 |
| SST-head motion, Mean (SD), mm | 0.39 (0.37) | 0.52 (0.48) * | 0.50 (0.50) * | >.05 |
| Cognition-Picture Vocabulary, Mean (SD) | 53.30 (11.17) | 50.22 (9.96) * | 50.43 (10.88) * | >.05 |
| Cognition-Flanker, Mean (SD) | 46.35 (9.04) | 46.42 (9.37) | 46.87 (9.47) | >.05 |
| Cognition-List Sorting, Mean (SD) | 50.35 (9.64) | 47.76 (10.35) * | 48.05 (10.03) * | >.05 |
| Cognition-Card Sort, Mean (SD) | 48.36 (9.88) | 46.70 (9.06) * | 46.72 (9.04) * | >.05 |
| Cognition-Pattern Comparison, Mean (SD) | 46.11 (14.00) | 44.61 (14.88) * | 45.40 (14.56) | >.05 |
| Cognition-Picture Sequence, Mean (SD) | 50.24 (11.26) | 49.35 (10.38) | 49.01 (10.30) | >.05 |
| Cognition-Oral Reading, Mean (SD) | 50.67 (11.90) | 45.96 (9.68) * | 46.89 (10.50) * | >.05 |
| Cognition-Fluid Composite, Mean (SD) | 47.00 (11.08) | 44.85 (11.35) * | 45.21 (10.88) * | >.05 |
| Cognition-Crystallized Composite, Mean (SD) | 52.35 (11.52) | 47.58 (9.39) * | 48.39 (10.35) * | >.05 |
| Cognition-Total Composite, Mean (SD) | 49.37 (11.14) | 45.04 (10.42) * | 45.74 (10.60) * | >.05 |
| Abbreviations: CBCL, Child Behavior Checklist; ELA, early-life adversity; SST, stop-signal task.  Group summaries describe demographic characteristics, ELA exposures, psychopathology, stop-signal task performance, head motion, and cognitive scores for the higher-income group and the 2 poverty subtypes. For continuous measures, between-group differences were evaluated using linear mixed-effects models with group (subtype) as the predictor, adjusting for baseline age, sex at birth, and race/ethnicity, and including random intercepts for site and family nested within site. For categorical measures, group differences were evaluated using generalized linear models with group (subtype) as the predictor and covariates aligned to the corresponding measure.  Asterisks indicate differences between each poverty subtype and the higher-income group that were significant after false discovery rate correction across measures (*P*_FDR_ < .05). *P* values in the rightmost column correspond to the direct comparison between subtype-1 and subtype-2; because no subtype 1 vs subtype 2 difference survived correction, these *P* values are reported without multiple-comparison adjustment. | | | | |

**Table 6. Differences in cumulative ELA–CBCL coupling between poverty neurofunctional subtype-1 and subtype-2 across waves**

|  | Δβ (95% CI) | | | |
| --- | --- | --- | --- | --- |
|  | Wave-1 (Baseline) | Wave-2 | Wave-3 | Wave-4 |
| Poverty subtype-1 vs higher-income | -0.140 (-0.207, -0.072) | -0.110 (-0.179, -0.041) | -0.111 (-0.181, -0.041) | -0.068 (-0.140, 0.003) |
| Poverty subtype-2 vs higher-income | -0.033 (-0.100, 0.034) | -0.076 (-0.144, -0.007) | -0.069 (-0.138, -0.000) | -0.062 (-0.134, 0.009) |
| Abbreviations: CBCL, Child Behavior Checklist; ELA, early-life adversity; β, standardized regression coefficient; CI, confidence interval. Entries show the difference in standardized cumulative ELA–CBCL slopes (Δβ = β_subtype-1/2_ – β_higher-income_) at each wave, based on the models described in Methods.  *, *P* < .05 after false discovery rate (FDR) correction across time-point comparisons. | | | | |

**Table 7. Regions showing significant (uncorrected) interactions with cumulative ELA and poverty status in predicting CBCL total** **problems**

|  | Prediction at wave-1 (baseline) | | Prediction at wave-2 | |
| --- | --- | --- | --- | --- |
| Brain Region | β (95% CI) | P value (uncorrected) | β (95% CI) | P value (uncorrected) |
| R-S collat transv ant | 0.04 (0.02 to 0.07) | 0.002 | 0.03 (-0.00 to 0.05) | 0.077 |
| L-G front inf-Orbital | 0.04 (0.01 to 0.06) | 0.010 | 0.02 (-0.00 to 0.05) | 0.091 |
| L-S orbital-H Shaped | 0.03 (0.01 to 0.06) | 0.017 | 0.02 (-0.01 to 0.05) | 0.225 |
| R-S circular insula ant | 0.03 (0.01 to 0.06) | 0.017 | 0.01 (-0.02 to 0.04) | 0.342 |
| L-S orbital med-olfact | 0.04 (0.01 to 0.07) | 0.021 | 0.03 (-0.00 to 0.06) | 0.075 |
| R-G Ins lg and S cent ins | 0.03 (0.00 to 0.06) | 0.022 | 0.02 (-0.01 to 0.05) | 0.117 |
| L-G orbital | 0.03 (0.00 to 0.06) | 0.034 | 0.02 (-0.01 to 0.05) | 0.159 |
| L-S temporal inf | -0.03 (-0.05 to -0.00) | 0.040 | -0.02 (-0.05 to 0.00) | 0.096 |
| L-G Ins lg and S cent ins | 0.03 (0.00 to 0.06) | 0.041 | 0.01 (-0.02 to 0.04) | 0.529 |

**Table 8. Moderation of cumulative-ELA-CBCL associations by inhibitory-control temperament and poverty/subtype status**

| Table 8.1 Association between cumulative ELA and CBCL total problems by poverty status and inhibitory-control temperament | | | | | | | |
| --- | --- | --- | --- | --- | --- | --- | --- |
|  |  | β (95% CI) | | | | | |
|  | Wave | Wave-3 | | | Wave-4 | | |
|  | Temperamental inhibitory control level | Low | Medium | High | Low | Medium | High |
| Higher-income group |  | 0.36 (0.28 to 0.44) | 0.29 (0.27 to 0.32) | 0.23 (0.19 to 0.27) | 0.34 (0.25 to 0.43) | 0.29 (0.26 to 0.32) | 0.25 (0.20 to 0.29) |
| Poverty-exposed group |  | 0.6 (0.45 to 0.75) | 0.41 (0.36 to 0.45) | 0.21 (0.11 to 0.31) | 0.61 (0.44 to 0.78) | 0.40 (0.34 to 0.45) | 0.18 (0.07 to 0.30) |
|  | Abbreviations: CBCL, Child Behavior Checklist; CI, confidence interval; ELA, early-life adversity; IC-L, low inhibitory-control temperament; IC-M, medium inhibitory-control temperament; IC-H, high inhibitory-control temperament; β, standardized regression coefficient.  Values represent standardized simple slopes (β) for the association between cumulative ELA and CBCL total problems within subgroups defined by poverty status and level of inhibitory-control temperament. Slopes were estimated using simple-slope analyses (emtrends in the emmeans package) from linear mixed-effects models including cumulative ELA, inhibitory-control temperament, poverty status, and their interactions, along with the covariates specified in the main Methods.  IC-L/IC-M/IC-H denote temperament values of **1/3/5** on the EATQ-R inhibitory-control subscale (range, 1–5). | | | | | | |

| Table 8.2 Association between cumulative ELA and CBCL total problems by neurofunctional subtype and inhibitory-control temperament | | | | | | | |
| --- | --- | --- | --- | --- | --- | --- | --- |
|  |  | β (95% CI) | | | | | |
|  | Wave | Wave-3 | | | Wave-4 | | |
|  | Temperamental inhibitory control level | Low | Medium | High | Low | Medium | High |
| Higher-income group |  | 0.35 (0.24 to 0.45) | 0.28 (0.25 to 0.32) | 0.22 (0.17 to 0.27) | 0.36 (0.24 to 0.47) | 0.30 (0.26 to 0.34) | 0.24 (0.19 to 0.30) |
| Poverty subtype-1 |  | 0.57 (0.26 to 0.88) | 0.42 (0.32 to 0.51) | 0.27 (0.08 to 0.46) | 0.49 (0.16 to 0.83) | 0.38 (0.27 to 0.48) | 0.26 (0.06 to 0.46) |
| Poverty subtype-2 |  | 0.83 (0.54 to 1.13) | 0.46 (0.37 to 0.56) | 0.09 (-0.09 to 0.26) | 0.83 (0.51 to 1.16) | 0.46 (0.35 to 0.56) | 0.08 (-0.11 to 0.27) |
|  | Abbreviations: CBCL, Child Behavior Checklist; CI, confidence interval; ELA, early-life adversity; IC-L, low inhibitory-control temperament; IC-M, medium inhibitory-control temperament; IC-H, high inhibitory-control temperament; β, standardized regression coefficient.  Values represent standardized simple slopes (β) for the association between cumulative ELA and CBCL total problems within subgroups defined by group/subtype (higher-income group, poverty subtype-1, and poverty subtype-2) and level of inhibitory-control temperament. Slopes were estimated using simple-slope analyses (emtrends in the emmeans package) from linear mixed-effects models including cumulative ELA, inhibitory-control temperament, group/subtype, and their interactions, plus the covariates described in the main Methods.  IC-L, IC-M, and IC-H again denote low, medium, and high levels of inhibitory-control temperament. | | | | | | |

**Table 9. Spatial correlations between subtype activation patterns and neurotransmitter receptor/transporter distributions**

|  | Activation map of poverty subtype-1 | | Activation map of poverty subtype-2 | | Study name |
| --- | --- | --- | --- | --- | --- |
| PET tracer / receptor/transporter | Correlation coefficient | *P* value | Correlation coefficient | *p* value |  |
| 5HT1a_1_ | .20 | .0483 | -.13 | >.05 | beliveau2017jneurosci |
| 5HT1a_2_ | .14 | >.05 | -.10 | >.05 | savli2012neuroimage |
| 5HT1b_1_ | -.08 | >.05 | .05 | >.05 | beliveau2017jneurosci |
| 5HT1b_2_ | -.06 | >.05 | .04 | >.05 | gallezot2010jcerebbloodflowmetab |
| 5HT1b_3_ | -.06 | >.05 | .04 | >.05 | savli2012neuroimage |
| 5HT2a_1_ | .08 | >.05 | -.02 | >.05 | savli2012neuroimage |
| 5HT2a_2_ | .03 | >.05 | -.01 | >.05 | beliveau2017jneurosci |
| 5HT2a_3_ | .10 | >.05 | .02 | >.05 | talbot2012neuroimage |
| 5HT4 | .14 | >.05 | -.06 | >.05 | beliveau2017jneurosci |
| 5HT6 | .02 | >.05 | .01 | >.05 | radhakrishnan2018jnuclmed |
| 5HTT_1_ | .09 | >.05 | .01 | >.05 | savli2012neuroimage |
| 5HTT_2_ | .10 | >.05 | -.04 | >.05 | beliveau2017jneurosci |
| CB1_1_ | -.07 | >.05 | .10 | >.05 | laurikainen2019neuroimage |
| CB1_2_ | .04 | >.05 | .03 | >.05 | normandin2015jcerebbloodflowmetab |
| D1 | .04 | >.05 | .01 | >.05 | kaller2017eurjnuclmedmolimaging |
| D2_1_ | .18 | .0499 | -.06 | >.05 | jaworska2020neuropsychopharmacology |
| D2_2_ | .17 | >.05 | -.05 | >.05 | smith2017jcerebbloodflowmetab |
| D2_3_ | .21 | .042 | -.09 | >.05 | sandiego2015jcerebbloodflowmetab |
| D2_4_ | -.03 | >.05 | -.04 | >.05 | alakurtti2015jcerebbloodflowmetab |
| DAT_1_ | .01 | >.05 | -.02 | >.05 | sasaki2012jnuclmed |
| DAT_2_ | .11 | >.05 | -.08 | >.05 | dukart2018scirep |
| GABAa | .11 | >.05 | .01 | >.05 | dukart2018scirep |
| GABAabz | -.06 | >.05 | .02 | >.05 | norgaard2021neuroimage |
| H3 | -.01 | >.05 | .07 | >.05 | gallezot2017jcerebbloodflowmetab |
| mGluR5_1_ | .01 | >.05 | .04 | >.05 | PI: Pedro Rosa |
| mGluR5_2_ | .06 | >.05 | .03 | >.05 | dubois2016eurjnuclmedmolimaging |
| mGluR5_3_ | -.01 | >.05 | .08 | >.05 | smart2019eurjnuclmedmolimaging |
| MOR_1_ | .05 | >.05 | .05 | >.05 | turtonen2021biolpsychiatrycnni |
| MOR_2_ | .00 | >.05 | .05 | >.05 | kantonen2020neuroimage |
| NET_1_ | .04 | >.05 | -.04 | >.05 | hesse2017eurjnuclmedmolimaging |
| NET_2_ | -.12 | >.05 | .02 | >.05 | ding2010synapse |
| VAChT_1_ | .03 | >.05 | -.01 | >.05 | PI: Lauri Tuominen & Synthia Guimond |
| VAChT_2_ | .08 | >.05 | -.05 | >.05 | bedard2019sleepmed |
| VAChT_3_ | -.02 | >.05 | .00 | >.05 | aghourian2017molpsychiatry |
| a4b2 | -.14 | >.05 | .05 | >.05 | hillmer2016neuroimage |
| M1 | .13 | >.05 | -.01 | >.05 | naganawa2020jnuclmed |
| Abbreviations: 5HT, 5-hydroxytryptamine; 5HTT, 5-hydroxytryptamine transporter; a4b2, α4β2 nicotinic receptor; CB1, cannabinoid receptor; D1, D2, dopamine receptors; DAT, dopamine transporter; GABA, γ-aminobutyric acid receptor; H3, histamine receptor; M1, muscarinic acetylcholine receptor; mGluR, metabotropic glutamate receptor; MOR, μ-opioid receptor; NET, norepinephrine transporter; PET, positron emission tomography; VAChT, vesicular acetylcholine transporter.  Entries show Pearson correlation coefficients between each PET-derived receptor/transporter distribution map and the subtype-specific activation map (poverty subtype-1 or poverty subtype-2; each contrasted against the higher-income group). P values were obtained using a spin-test procedure with 10,000 permutations to account for spatial autocorrelation. Study names correspond to the original PET datasets summarized in Hansen et al. (2022). P values are not corrected for multiple comparisons and should be interpreted as exploratory. | | | | | |
